# Effects of the DailyColors™ Polyphenol Supplement on Serum Proteome, Cognitive Function, and Health in Older Adults at Risk of Cognitive and Functional Decline

**DOI:** 10.1101/2024.12.06.24318594

**Authors:** Mary O’Leary, Joanna L Bowtell, Megan Richards, Esra Bozbaş, Abbie Palmer, Kate Stych, Monica Meng, Adam Bloomfield, Lauren Struszczak, Jack Pritchard, Celeste Lugtmeijer, George Vere, Raif Yücel, Ana Rodriguez Mateos, Zicheng Zhang, Jonathan CY Tang, Clive Ballard, Anne Corbett

## Abstract

The Mediterranean diet is associated with reduced mortality and cognitive decline, largely due to its polyphenol content. However, Western populations often do not meet recommended fruit and vegetable intakes. Polyphenols exert anti-inflammatory effects and may influence extracellular vesicle (EV) dynamics. DailyColors™ is a polyphenol-rich blend inspired by the Mediterranean Diet, containing extracts from 16 fruits, vegetables, and herbs. This 60-day, double-blind, placebo-controlled, randomised trial involved 150 UK adults aged 50+ with a BMI ≥ 25, recruited to complete cognitive and physical fitness assessments via the PROTECT-UK online platform. Participants received either a medium (750mg) or high (2000mg) dose of DailyColors™ (∼300mg and ∼750mg polyphenols, respectively) or a placebo. A sub-group (n=15 per group) underwent additional assessments, including blood pressure measurements, characterisation of circulating EVs and tandem-mass-tagged serum proteomics. Significant cognitive benefits were observed, with improvements in reaction time for the high-dose group and accuracy for both active supplement groups. The high-dose group also showed significant physical fitness gains on the Timed Stand Test (P<0.001). All groups improved on the Chair Stand Test. Proteomic analysis showed significantly reduced serum protein expression in immune and pre-β1-HDL pathways, suggesting anti-inflammatory effects. Pre-β1-HDL proteins are typically elevated in obesity; their reduction suggests a reversal of this obesity-related effect. No significant changes were noted in EV concentration or size. DailyColors™ supplementation, may enhance cognitive function, physical fitness, and systemic health in older, overweight adults, potentially mimicking the anti-inflammatory effects of the Mediterranean diet. These findings warrant investigation in larger trials.

## Introduction

Consumption of a diet rich in fruits and vegetables, such as the Mediterranean diet, is associated with a reduced risk of all-cause mortality, especially from cardiovascular disease, and a slower rate of cognitive decline with age [1–4]. However, adopting the Mediterranean diet outside of its native region can be challenging, with financial barriers, limited ingredient accessibility, nutritional knowledge gaps, and other socio-cultural factors impacting uptake [5]. Less than a third of British adults consume the recommended five portions of fruit and vegetables per day—an intake level linked to reduced morbidity and mortality [3,6]. Epidemiological evidence suggests that polyphenols within these foods play a key role in reducing cardiovascular disease risk and cognitive decline [7,8]. Despite the health advantages, achieving population-wide increases in fruit, vegetable, and polyphenol intake remains difficult. Therefore, there is a pressing need for alternative strategies with low barriers to adoption that deliver similar health benefits.

Dietary polyphenols appear to exert their effects primarily through anti-inflammatory and antioxidant mechanisms. Cross-sectional human studies reveal that individuals with higher polyphenol intakes generally show reduced levels of inflammation [9], supporting the observed links to improved vascular [10–12] and cognitive [13–15] health. Interventional studies in animal models offer further evidence, showing that polyphenols can significantly lower pro-inflammatory cytokines [16–19], enhance antioxidant defences [20,21], and improve cognition [22–25]. Additionally, polyphenols may improve endothelial function and reduce atherosclerosis in animal models—both crucial factors in cardiovascular health [26].

Interventional human studies reinforce these findings, with randomised controlled trials showing decreases in inflammation among those consuming polyphenol-rich foods or supplements [27–30]. Human trials also suggest cognitive, metabolic and vascular benefits, [31–35]

There is emerging evidence of mechanisms of polyphenol action that appear to represent extensions of the well-established anti-inflammatory and antioxidant processes, offering a deeper understanding of how these actions translate into functional health effects. These include epigenetic modifications and alterations in extracellular vesicles [36,37]. Extracellular vesicles (EVs) are released by various cell types in response to cellular stress, activation, and apoptosis, and carry a cargo of bioactive molecules including phenolic compounds. They are suggested to play an important role in cell-to-cell communication and are associated with different clinical conditions and aging [37]. A Mediterranean diet intervention elicited a reduction in the number of some EVs among elderly participants [38], and reduced prothrombotic vesicle release in individuals at high cardiovascular disease risk [39]. Cocoa flavanol supplementation has also been shown to decrease circulating endothelial derived EVs in overweight young participants (700 mg.d^-1^ for 4 weeks) [40] and in elderly participants (450 mg.d^-1^ for 2 weeks) [41]. These complex emerging mechanisms provide a strong argument for hypothesis-free ‘omic’ analyses of tissues in those supplemented with polyphenols. Such analyses could provide a more comprehensive understanding of their effects across various biological systems.

Improving cognitive health trajectory in older adults is a critical public health issue since early cognitive impairment is a major risk factor for dementia, a devastating condition which affects one million people in the UK [42]. Several medical and lifestyle factors are known to increase risk of dementia, and 40% of dementia cases are potentially preventable through addressing modifiable risk [43]. Older adults are increasingly engaged with the importance of maintaining cognitive health and research shows a desire for accessible interventions to achieve this [44]. Dietary supplements have been raised as a potential means of benefitting cognitive health in combination with other lifestyle interventions such as cognitive training, yet evidence is mixed regarding efficacy of supplements to elicit change [45]. Epidemiological evidence supporting the benefits of the Mediterranean diet is strong but implementation of a dietary intervention is challenging [46].

DailyColors^TM^ is a patented proprietary blend of extracts and freeze-dried powders of 16 fruit, vegetables and herbs, inspired by the Mediterranean Diet and rich in polyphenols. Previous analyses have suggested that DailyColors^TM^ contains a broad range of bioactives, including phenolics, flavonoids, anthocyanins and chlorogenic acids. One month of DailyColors^TM^ supplementation has been observed to increase hypermethylation of the candidate CpG site cg13108341, contrary to the trend observed at this site during ageing [47]. However, the ability of DailyColors^TM^ to mimic the health effects of the Mediterranean Diet and to reflect known health effects of dietary polyphenols must be interrogated further.

This study aimed to determine the physiological and cognitive effects of 60 days of supplementation with either a low (750 mg.d^-1^) or high (2000 mg.d^-1^) dose of the DailyColors^TM^ supplement versus a placebo in overweight older adults. The UK traditional diet provides approximately 1000mg.d^-1^ polyphenols [48]. We interrogated the effects of supplementation on cognition digital measures of health and wellbeing, blood pressure, the serum proteome and extracellular vesicles.

## Experimental Methods

### Study Design

This was a 60-day mechanistic three-arm double-blind placebo-controlled randomised clinical trial delivered through a remote and hybrid design using the PROTECT-UK research platform [49]. The study received ethical approval from the University of Exeter Public Health & Sports Sciences Research Ethics Committee (Ref: 2929219). This trial was retrospectively registered: ISRCTN10734674

### Participants

Participants were 150 adults aged 50 and above who were already participants on the PROTECT-UK ageing cohort study and residing in the UK. Eligible participants had a Body Mass Index of 25 or above and living within two hours’ travel time from Exeter. Participants were ineligible if they had a vegan diet, consumed more than five servings of coffee per day or regularly consumed anti-inflammatory medications, polyphenol-rich dietary supplements or medications contraindicated for high doses of grapefruit. The first 45 participants enrolled in the study were invited to complete additional physiological and proteomic assessments. The remainder of the cohort completed the trial remotely from home.

### Recruitment

Participants were recruited from the PROTECT-UK cohort using the cohort’s consent-for-contact protocol. Pre-screening of existing data was performed to identify individuals fulfilling the age, location and BMI eligibility criteria, and participants were contacted by email to register for the study in a bespoke area of the PROTECT-UK website. Participants completed a screening questionnaire for all eligibility criteria prior to giving informed consent using an ethically approved digital process. The sub-group of participants for in-person assessments completed a second consent process for the additional activities using a written informed consent process.

### Treatment Interventions

Participants were allocated to one of three treatment groups. Active treatment groups received a tablet containing the DailyColors^TM^ phytonutrient supplement packed into microcrystalline cellulose capsules containing additional inert packing ingredients of Natural Rice Concentrate (NuFlow) and Natural Rice Extract (NuMag) at quantities to ensure the correct dose. The low dose group received 187.5mg DailyColors^TM^ per tablet, and the high dose group received 500mg DailyColors^TM^ per tablet. The placebo capsules consisted of identical capsule and inert packing components and no DailyColors^TM^ components (See Supplementary Table 1). Participants received their tablets in the post or at clinic visits if they were recruited to the sub-group assessment protocol and were instructed to take two tablets in the morning and two in the evening for a 60-day period.

### Randomisation and Masking

Randomisation of participants was achieved through minimisation randomisation using a validated randomisation software. This occurred after a participant had consented to take part in the trial. The randomisation algorithm stratified participants by age (age brackets of five years), sex and BMI (brackets of 25 – 29.9, 30-39.9, 40+). Treatment allocation was recorded electronically by an independent research staff member. Both participants and research team were blind to allocation, thus removing any bias. Administrative staff on the trial helpdesk that had direct participant contact were not involved in any data analysis to avoid any unconscious bias. Similarly, the web developers involved in maintenance of the trial infrastructure were not involved in any data analysis or interpretation.

### Supplement Analysis

The high dose participants consumed a daily (poly)phenol dose of 462 mg.d^-1^ (94 mg anthocyanins, 9 mg flavonols, 109 mg phenolic acids, 180 mg flavon-3-ols, and 70 mg other (poly)phenols), whilst low dose participants received 247 mg.d^-1^ (34 mg anthocyanins, 5 mg flavonols, 67 mg phenolic acids, 87 mg flavon-3-ols, and 53 mg other (poly)phenols). Full details of the supplement composition and analytical methods are available in Supplementary Tables 2 and 3.

Outcome measures: The full cohort completed measures of cognition, health and wellbeing through the PROTECT-UK platform. Cognition was assessed using the FLAME cognitive test battery, which is validated to detect pre-clinical change in working memory, episodic memory, attention and executive function [50]. Health and wellbeing were assessed using the Patient Health Questionnaire (PHQ-9) for depression [51], General Anxiety Disorder scale (GAD-7) for anxiety [52], Instrumental Activities of Daily Living scale for function [53], IQCODE for subjective cognitive performance [54], an adapted sleep questionnaire and EQ-5D for wellbeing [55]. Physical fitness was measured remotely using a battery of well-established clinical assessments including the Two-Minute Step Test [56], Timed Chair Stand test [57], Chair-Stand test and self-reported gait velocity.

For the sub-group, further assessments were performed in-person at a clinical facility at baseline and after 60 days of supplementation. Following 5 minutes of rest in a seated position, systolic and diastolic blood pressure were measured three times using a sphygmomanometer. Body weight and height were recorded. Venous blood samples were collected from overnight fasted participants into vacutainer tubes (serum silica tubes, plasma lithium heparin tubes, EVs 3.2% sodium citrate tubes; Greiner Bio-One, UK) through a large diameter, 21-gauge needle mounted on a 19cm length of plastic tubing (BD Medical, USA) at each visit. No tourniquet was used during collection of samples for downstream EV analyses. Two samples were collected at each visit, a basal sample upon arrival at the facility and a further sample was collected 2h after ingestion of the first (day 1) and last (day 60) dose of supplement.

Sample size: This was an exploratory trial to determine the mechanistic action of a dietary supplement and to collate data of indicative effectiveness across a broad range of cognitive, health genetic and physiological outcomes. The sample size is therefore based on a pragmatic trial design to allow for collection of the outcome measures. An indicative power calculation for the cognitive outcomes was based on existing trials of polyphenols with cognitive outcomes, which showed that 150 participants would be required to detect an effect size of 0.3 at a significance level of 0.05 with 45% power, allowing for 20% dropout (23). The subgroup sample size of 15 was determined based on our previously published skeletal muscle tissue proteomic analyses [58].

### Safety Monitoring

Safety monitoring was achieved through participant report of adverse events using an online report function on the PROTECT-UK dashboard for this trial. Participants were able to access an online questionnaire that captured data required for evaluation of any adverse events, which prompted review by the trial management team and reporting as appropriate for Good Clinical Practice. Participants were also able to contact the study helpdesk to report an adverse event at any time during the study.

### Cognitive Data Analysis

Cognitive data was processed to provide total scores for individual tests and cognitive domains, and subject to a pre-post analysis using paired t-tests, controlling for age and sex. Data for health and wellbeing data was also analysed using this methodology.

### Phenolic Blood Analysis

Plasma samples were analysed for phenolic metabolite concentration at the Bioanalytical Facility, University of East Anglia. Plasma concentrations of protocatechuic acid, 4-hydroxybenzoic acid, hippuric acid, vanillic acid, ferulic acid and isoferulic acid were quantified using a Waters Xevo TQ-XS tandem mass spectrometer coupled with an Acquity I-class ultra high-pressure liquid chromatography pump (UPLC) system (Waters Corp., Milford, MA, USA) as described previously [59]. The plasma sample extraction procedure was described in [60]and detailed here in the supplementary materials with the assay performance statistics (Supplementary Table 4).

### Extracellular Vesicle Blood Analysis

To obtain platelet-free plasma (PFP), venous blood was centrifuged at 2,500 x g for 15 minutes, with no brake at room temperature and the upper two-thirds were collected and centrifuged again at 2,500 x g for 15 minutes, with no brake at room temperature. The upper three-quarters of each tube were collected and identified as PFP. Prepared PFP was aliquoted and stored at −80 °C for further analysis.

### Isolation

Size exclusion columns (qEVoriginal Gen 2, Izon Science) were used to isolate circulating EVs from PFP as recommended by the manufacturer. The column within the operational temperature range (18-24°C) was equilibrated with 30 ml of PBS (Sigma-Aldrich, Dorset, UK), then 0.5 ml of PFP was loaded on top of the luer-slip cap and allowed to enter the column. The top reservoir was then filled with PBS and the first 2.5 ml of eluent was collected in separate collection tubes as these initial fractions primarily contain larger proteins and debris. Then 0.4 ml of eluent was collected per fraction and the EV-rich fractions (2∼4) were obtained and pooled together.

### Nanoparticle Tracking Analysis

The size distribution and concentration of EVs were determined by Nanoparticle Tracking Analysis (NTA) using a NanoSight 300 (NS300; Malvern, Amesbury, UK). Isolated EV samples by SEC were diluted in filtered, sterile PBS (Sigma-Aldrich, Dorset, UK) to carry out the measurement and manually injected into the NTA sample chamber using a 1 ml syringe and syringe pump at room temperature. Three 1-minute videos of each diluted sample were captured at camera level 13 and frame rate of 25 per second and analysed by Nano 3.4 software. The isolation of EVs using SEC results in highly purified EVs, but some lipoprotein classes may co-isolate with them [61]; in order to achieve the best possible separation from lipoproteins, a threshold of 70 nm diameter was set. Since the majority of lipoproteins are smaller than this and the samples were always from fasted subjects, the EV preparations were essentially free of lipoprotein contamination. However, the threshold of 70 nm meant that EVs smaller than this could not be captured.

### Flow cytometry

In this study, appropriate reporting methods followed MIFlowCyt guidelines, detailing the flow cytometer settings and reagents used [1]. Prior to flow cytometry, 5 μl of isolated EVs were stained with 40 μM CFSE (ThermoFisher Scientific, Paisley, UK), then an antibody staining cocktail of anti-CD31-PE (BD Bioscience, Oxford, UK), anti-CD41a-APC (BD Bioscience, Oxford, UK), and anti-CD45-BV711 (BD Bioscience, Oxford, UK). Stained EVs were diluted in filtered PBS before acquisition on a 5-laser Cytek Aurora Flow Cytometer with an enhanced small particle detection module (ESP) using SpectroFlo software (Cytek Biosciences Inc, Fremont, CA). SSC and fluorescence gains were increased to the resolution of EVs and a threshold for both SSC/BL1 (SSC:750 OR BL1:750) was set. Samples were run on a low flow rate for 2 minutes. Sizing of EVs was conducted using Mie scatter interpolation via FCMPASS (Welsh 2019 Cytometry A) calibrated for each measurement using ApogeeMix beads (Cat#1527, Apogee Flow Systems, UK). FCMPASS-processed FCS files were then gated in FlowJo v10.10.0 (BD Life Sciences) to identify EV populations using fluorescence minus one (FMO) controls to gate total (CFSE^+^), platelet-derived EVs (CD41^+^) and endothelial cell-derived EVs (CD31^+^CD45^-^) and immune-derived (CD45^+^) EVs (Supplementary Figure 1A and B), and size/concentration of these populations exported for downstream analysis [62,63]. Detection of coincident EV events (swarming) was ruled out by performing serial dilution of isolated EVs which showed a linear decrease in event rate (Supplementary Figure 1C). This corresponded to approximately 5000 events per second, and participant samples were diluted to run at this event rate.

### Proteomics

Serum samples (10ul) were depleted of the 14 most abundant proteins using High-Select™ Top14 Abundant Protein Depletion Resin, according to the manufacturer’s protocol (Pierce). Aliquots of 30µg of each depleted sample were digested with trypsin (1.25µg trypsin; 37°C, overnight), labelled with Tandem Mass Tag (TMTpro) eighteen plex reagents according to the manufacturer’s protocol (Thermo Fisher Scientific, Loughborough, LE11 5RG, UK) and the labelled samples pooled.

An aliquot of 200ug of the pooled sample was desalted using a SepPak cartridge according to the manufacturer’s instructions (Waters, Milford, Massachusetts, USA). Eluate from the SepPak cartridge was evaporated to dryness and resuspended in buffer A (20 mM ammonium hydroxide, pH 10) prior to fractionation by high pH reversed-phase chromatography using an Ultimate 3000 liquid chromatography system (Thermo Fisher Scientific). In brief, the sample was loaded onto an XBridge BEH C18 Column (130Å, 3.5 µm, 2.1 mm X 150 mm, Waters, UK) in buffer A and peptides eluted with an increasing gradient of buffer B (20 mM Ammonium Hydroxide in acetonitrile, pH 10) from 0-95% over 60 minutes. The resulting fractions (concatenated into 20 in total) were evaporated to dryness and resuspended in 1% formic acid prior to analysis by nano-LC MSMS using an Orbitrap Fusion Lumos mass spectrometer (Thermo Scientific).

High pH RP fractions were further fractionated using an Ultimate 3000 nano-LC system in line with an Orbitrap Fusion Lumos mass spectrometer (Thermo Scientific). In brief, peptides in 1% (vol/vol) formic acid were injected onto an Acclaim PepMap C18 nano-trap column (Thermo Scientific). After washing with 0.5% (vol/vol) acetonitrile 0.1% (vol/vol) formic acid peptides were resolved on a 250 mm × 75 μm Acclaim PepMap C18 reverse phase analytical column (Thermo Scientific) over a 150 min organic gradient, using 7 gradient segments (1-6% solvent B over 1min., 6-15% B over 58min., 15-32%B over 58min., 32-40%B over 5min., 40-90%B over 1min., held at 90%B for 6min and then reduced to 1%B over 1min.) with a flow rate of 300 nl min−1. Solvent A was 0.1% formic acid and Solvent B was aqueous 80% acetonitrile in 0.1% formic acid. Peptides were ionized by nano-electrospray ionization at 2.0kV using a stainless-steel emitter with an internal diameter of 30 μm (Thermo Scientific) and a capillary temperature of 300°C.

All spectra were acquired using an Orbitrap Fusion Lumos mass spectrometer controlled by Xcalibur 3.0 software (Thermo Scientific) and operated in data-dependent acquisition mode using an SPS-MS3 workflow. FTMS1 spectra were collected at a resolution of 120 000, with an automatic gain control (AGC) target of 200 000 and a max injection time of 50ms. Precursors were filtered with an intensity threshold of 5000, according to charge state (to include charge states 2-7) and with monoisotopic peak determination set to Peptide. Previously interrogated precursors were excluded using a dynamic window (60s +/-10ppm). The MS2 precursors were isolated with a quadrupole isolation window of 0.7m/z. ITMS2 spectra were collected with an AGC target of 10 000, max injection time of 70ms and CID collision energy of 35%.

For FTMS3 analysis, the Orbitrap was operated at 50 000 resolution with an AGC target of 50 000 and a max injection time of 105ms. Precursors were fragmented by high energy collision dissociation (HCD) at a normalised collision energy of 60% to ensure maximal TMT reporter ion yield. Synchronous Precursor Selection (SPS) was enabled to include up to 10 MS2 fragment ions in the FTMS3 scan.

The raw data files were processed and quantified using Proteome Discoverer software v2.4 (Thermo Scientific) and searched against the UniProt Human database (downloaded January 2024: 82415 entries) using the SEQUEST HT algorithm. Peptide precursor mass tolerance was set at 10ppm, and MS/MS tolerance was set at 0.6Da. Search criteria included oxidation of methionine (+15.995Da), acetylation of the protein N-terminus (+42.011Da) and Methionine loss plus acetylation of the protein N-terminus (−89.03Da) as variable modifications and carbamidomethylation of cysteine (+57.0214) and the addition of the TMTpro mass tag (+304.207) to peptide N-termini and lysine as fixed modifications. Searches were performed with full tryptic digestion and a maximum of 2 missed cleavages were allowed. The reverse database search option was enabled and all data was filtered to satisfy false discovery rate (FDR) of 5%.

Data were analysed at the University of Exeter. Data were normalised to the total peptide amount in each sample and scaled using a pooled ‘reference’ sample common to both TMT experiments to facilitate the comparison of protein levels between experiments. Data were log_2_ transformed prior to analysis. Data were filtered to include only proteins that were detected in all samples (432 proteins). Reactome (Reactome V8 (https://reactome.org/)) was used to perform differential gene expression analysis using the ‘Correlation Adjusted MEan RAnk gene set test’ (CAMERA) algorithm. The false discovery rate (FDR, Benjamini-Hochberg-adjusted p-value < 0.05) and log_2_ fold change (log_2_FC) values were calculated by Reactome. Weighted Gene Co-expression Network Analysis (WGCNA) was performed to identify co-expressed protein modules in the whole dataset (R version 4.3.2, WGCNA version 1.72-1. A power parameter with a signed R2 above 0.80 was chosen, with a soft threshold of 10 used for WGCNA). Module eigengenes were correlated with physiological traits using WGCNA’s weighted Pearson correlation. Student’s asymptotic p-value was calculated for these correlations. Correlations between physiological traits were calculated using the same approach.

### Other Statistical Analysis

Other data (plasma phenolics, EVs, physiological measures) were analyzed using ANCOVA, with pre-supplementation baseline values as a covariate. Analysis was conducted in IBM SPSS Statistics v29.

## Results

### Cohort Characteristics

150 participants were recruited to this study, of whom 45 were recruited to the sub-group for physiological and proteomic analysis. The mean age of participants was 64 (SD 6.7), 80% of the cohort was female and the mean BMI was 30 (SD 5.09) kg.m^-2^. The three intervention groups were well balanced for the stratification factors of age, sex and BMI with no between group differences at baseline (Supplementary Table 5). Of these, 141 completed the 60-day study in the full cohort and 37 completed the sub-group analysis (Figure 1).

**Figure 1.**
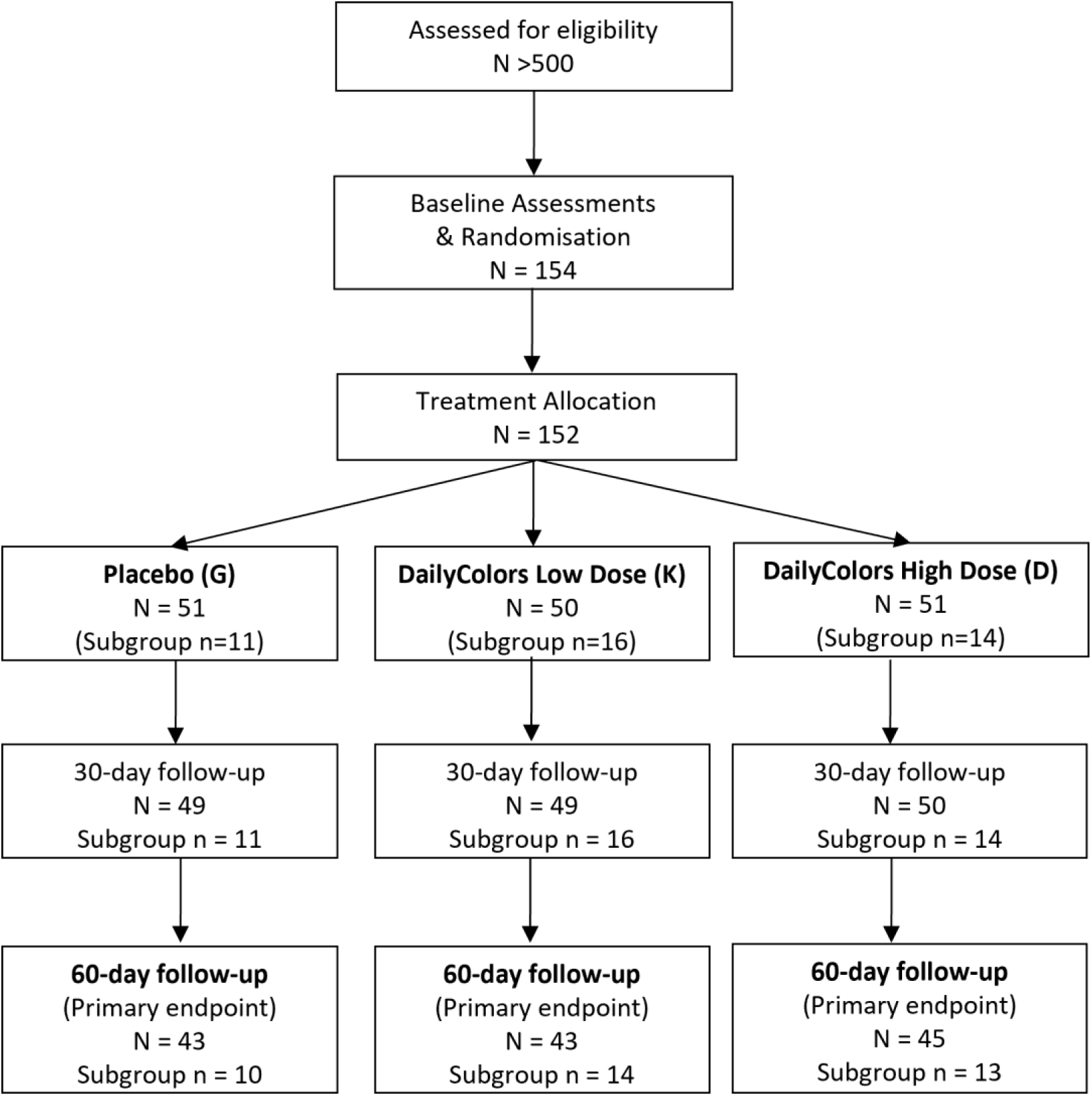
CONSORT Chart showing flow of participants through the study.

### Anthropometric Data and Blood Pressure

There was no effect of supplementation on anthropometric data. There was a significant condition effect for systolic blood pressure (p=0.024) when controlled for baseline, with a greater increase in SBP in the high versus low dose condition (p=0.023) after 60d supplementation.

### Effect of DailyColors^TM^ supplementation on cognition and digital measures of health and wellbeing

Analysis of cognitive data showed a significant benefit to attentional measures of reaction time (Digit Vigilance: Cohen’s D effect size 0.34, P=0.042) in the high dose group and accuracy in both active intervention groups (Delayed Picture Recognition: Low Dose group ES = 0.41, P = 0.015; High Dose group ES = 0.42, P = 0.014) but not in the placebo group. No significant impact was seen on measures of memory or executive function.

For online measures of physical fitness, significant improvement was seen in the High Dose group in the Timed Stand Test (P<0.001) and on the Chair Stand Test in all groups, with the greatest impact on the active intervention groups (P<0.001). In wellbeing measures, significant improvement was seen across all groups for depression (P<0.0001) but no impact was seen on other wellbeing measures (Supplementary Table 6).

### Phenolic Metabolites

Sixty days of supplementation did not alter plasma concentration of any of the measured phenolic metabolites in the fasted state. However, there was a significant elevation of some of the measured plasma phenolic metabolite concentrations 2 h after ingestion of the first dose (isoferulic, protocatechuic and vanillic acids) and final dose (protocatechuic and vanillic acids) of supplement; whilst plasma hippuric acid concentrations decreased 2 h after ingestion of first and final supplement dose (Figure 2). Two hours after ingestion of the first and final supplement doses, plasma ferulic (P<0.05) and vanillic acid (P<0.001) concentrations were significantly higher in the high dose condition than both placebo and Low Dose conditions, when controlled for baseline concentrations. Additionally, 2 hours after the final dose, plasma concentrations of isoferulic acid (P<0.05) and vanillic acid (P<0.001) were significantly higher in the Low Dose group compared to the placebo, after adjusting for baseline levels. In contrast, 2 hours after the final dose, plasma hippuric acid concentration was significantly lower in the Low Dose group compared to the placebo, after adjusting for baseline levels (P<0.005).

**Figure 2.**
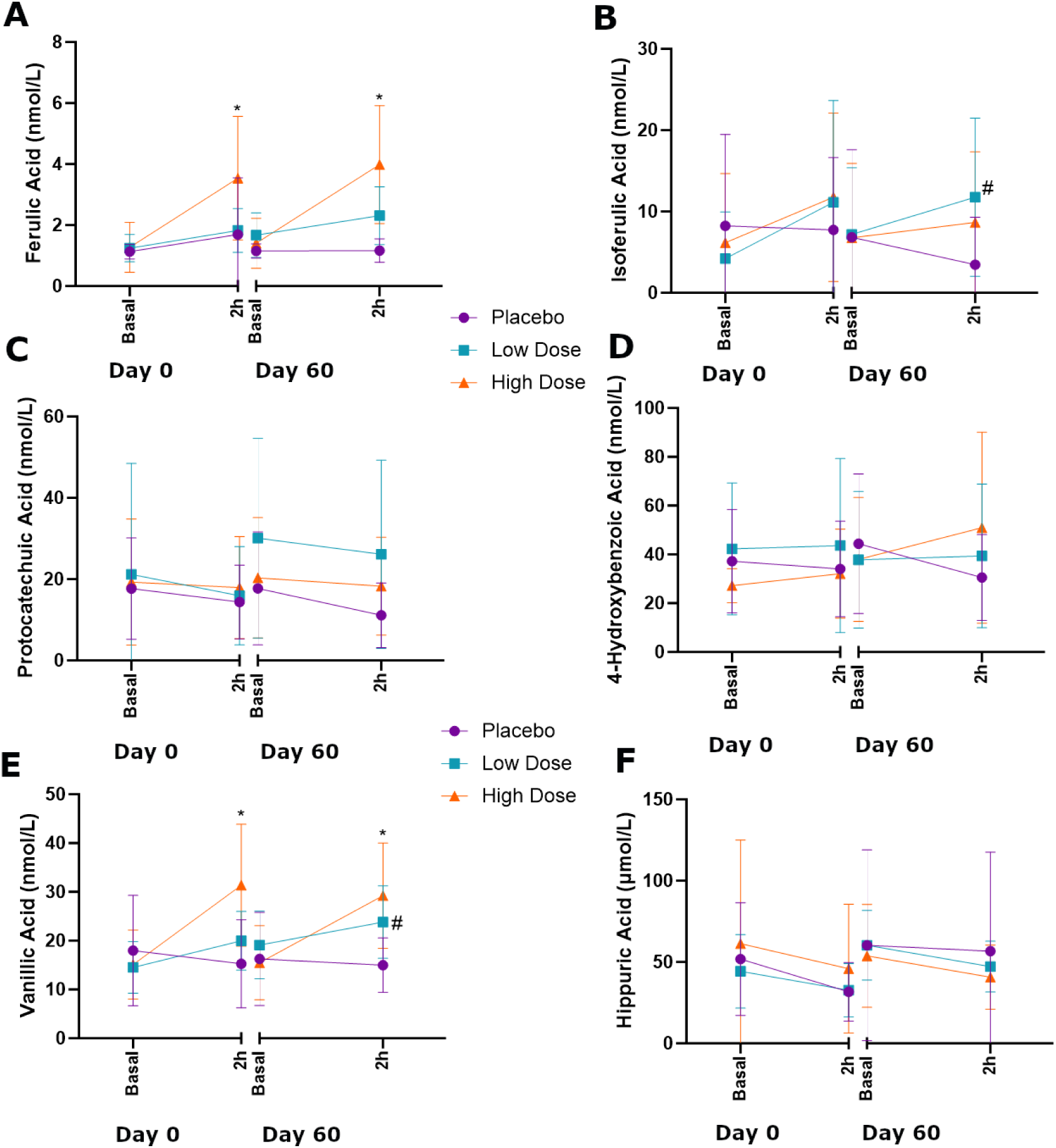
Effect of sixty days DailyColors™ supplementation on plasma phenolic metabolite concentrations. Plasma concentrations of phenolic metabolites were measured in the fasted state and 2 hours after ingestion of the first and final supplement doses. DailyColors™ High Dose = 2000 mg DailyColors™. DailyColors™ Low Dose = 750 mg DailyColors™ combined with microcrystalline cellulose as a filler. Placebo = microcrystalline cellulose.

### Extracellular Vesicles

The DailyColors™ supplementation had no significant effect on total EV concentration/size or the size/concentration of specific EV populations (CD31^+^, CD41 ^+^, CD45 ^+^, or CD31^+^CD45^-^), following the 60-day intervention (Supplementary Figures 2-4).

### Effect of DailyColors™ Supplementation on the Serum Proteome

Sixty days dietary supplementation with DailyColors™ significantly reduced the expression of proteins in pathways related to the adaptive immune system, the innate immune system, haemostasis, high-density lipoprotein assembly and vesicle-mediated transport. High-dose and low-dose DC treatments elicited congruent effects across multiple pathways (Table 1). These effects were distinct from those of the placebo, with no overlap in the serum proteomic responses. The placebo reduced the expression of proteins in pathways related to chromatin organisation, smooth muscle contraction, protein folding, epigenetic regulation of gene expression and DNA repair (Table 1).

**Table 1.**
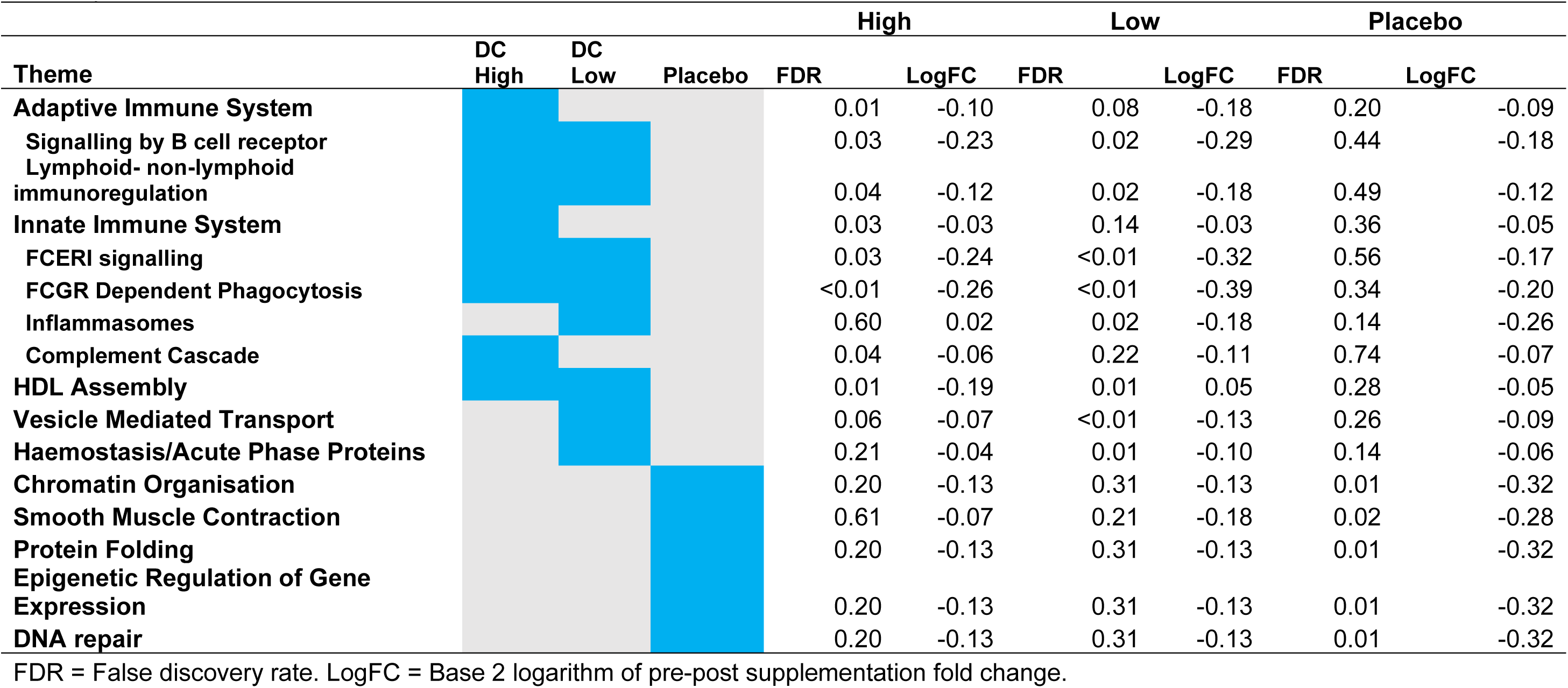
Proteomic Themes Differentially Regulated by 60 Days DailyColours or Placebo Supplementation. Reactome parent and first-level child pathways that significantly differed following supplementation with DailyColors ‘High dose’ (2000 mg) ‘Low dose’ (750 mg) or ‘Placebo’ (microcrystalline cellulose) are presented. Second level immune-related pathways are presented for completeness. Disease-related

WGCNA identified two protein modules. Module 1 contained 23 – largely cytoskeletal – proteins (Supplementary File 2). For these analyses, supplementation status was coded as ‘0’ (placebo) or ‘1’ (DailyColors™, either dose). Module 1 was not significantly correlated with supplementation status (DailyColors™ vs placebo). However, this module displayed significant correlations with systolic blood pressure (r = 0.29, p = 0.020), balance (r = −0.2, p = 0.026), attention (r = −0.30, p = 0.018), serum isoferulic acid 2h following supplement consumption (r = −0.25, p = 0.047), and baseline serum 4-hydroxybenzoic acid (r = −0.30, p = 0.017).

Most (406) other detected – largely acute phase – serum proteins were grouped into Module 0. Proteins in this module were significantly correlated with supplementation status; those supplemented with DC displayed significantly lower serum concentrations of Module 0 proteins compared to placebo supplemented individuals (r = −0.30, p =0.015).

Correlations between physiological traits were calculated. Supplementation with DailyColors was positively correlated with ferulic acid and isoferulic acid concentrations 2h post supplementation (Figure 3); corroborating data presented in Figure 1. DailyColors supplementation was positively correlated with measures of improved cognitive function (executive function, picture recognition accuracy, numerical working memory). DailyColors supplementation was significantly correlated with higher diastolic blood pressure (Figure 3).

**Figure 3.**
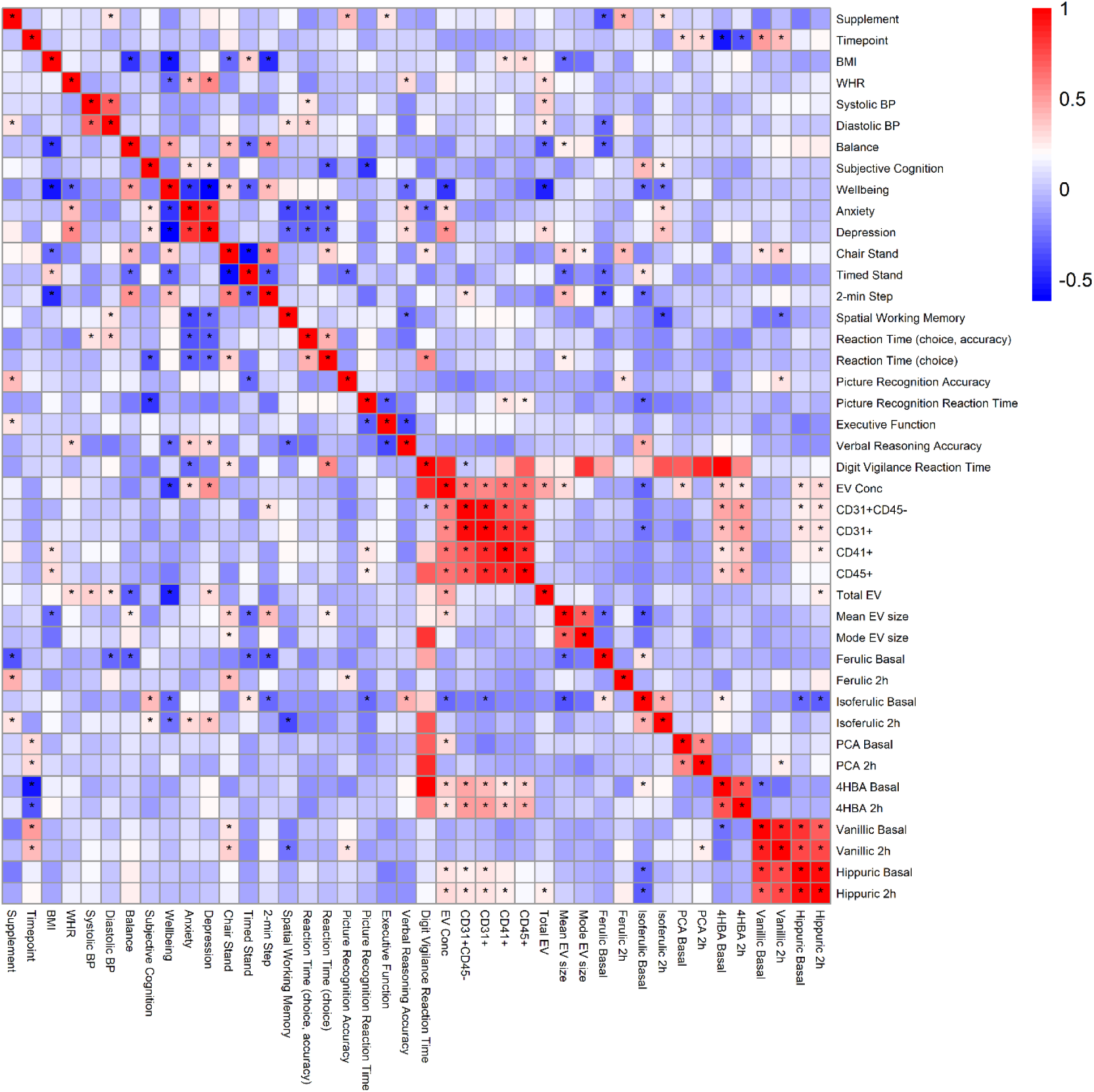
Pearson correlations between physiological traits in participants supplemented with DailyColors ‘High dose’ (2000 mg) ‘Low dose’ (750 mg) or ‘Placebo’ (microcrystalline cellulose) for 60 days. For these analyses, supplementation status was coded as ‘0’ (placebo) or ‘1’ (Dailycolors, either dose) A blue−white−red gradient indicates Pearson value. * = p <0.05 for Pearson correlation. EV conc = total extracellular vesicle concentration, flow cytometry. CD31^+^, CD41 ^+^, CD45 ^+^, CD31^+^CD45^-^ denote concentrations of specific EV populations. Total EV, mean EV size and mode EV size measured via nanoparticle tracking analysis. Basal = average of data taken before first or final dose of supplement. 2h = average of data taken 2 hours following first or final dose of supplement.

Positive correlations were observed between EV surface markers and plasma 4HBA and hippurate concentrations. CD41+ and CD45+ EV concentrations were positively correlated with picture recognition reaction time and BMI. Total EV numbers were positively correlated with higher systolic and diastolic blood pressure. Mean EV size was positively correlated with better physical performance and balance scores and was negatively correlated with BMI (Figure 3).

Lower depression and anxiety scores were significantly correlated with better cognitive function (spatial working memory and choice reaction time; Figure 3).

## Discussion

This study investigated the effects of 60 days of DailyColors™ supplementation on physiological, cognitive, and proteomic outcomes in an older adult cohort. Significant improvements were observed in cognitive function, particularly in attentional measures and reaction time, as well as in physical fitness, with marked benefits noted in the high-dose group. Serum proteomic analysis revealed that DailyColors™ elicited changes in protein expression in pathways related to inflammation and immunity, vesicle-mediated transport, and high density lipoprotein (HDL) assembly, distinct from those observed in the placebo group. Collectively, these findings provide early evidence that DailyColors™ may confer cognitive and systemic health benefits, with dose-responsive effects evident for cognitive measures and physical fitness outcomes.

The significant improvements in attentional reaction time and accuracy in the DailyColors™ groups align with prior findings that attentional measures often show early responsiveness to nutritional interventions. Furthermore, despite being underpowered to provide a definitive outcome, the effect sizes achieved for these attentional measures are substantial and are within the same range as those reported for pharmacological interventions in the cognitive health space [64,65]. This further indicates promising evidence of cognitive benefit. As attention and reaction time are sensitive markers of cognitive health, these findings underscore the potential of DailyColors™ supplementation as an accessible approach for enhancing early-stage cognitive function in aging populations.

Physical fitness improvements, such as the significant gains in the Timed Stand Test in the high-dose group and in the Chair Stand Test across all groups, support the notion that DailyColors™ may bolster global physical function. These outcomes could reflect underlying physiological enhancements, possibly through modulation of inflammatory or metabolic pathways, as indicated by proteomic findings; these are considered below. The general improvement in depression scores across all groups, including placebo, likely represents a broader cohort effect, suggesting that engagement in the study itself may have positively impacted participants’ mental wellbeing.

DailyColors™ supplementation significantly downregulated proteins involved in both adaptive and innate immune pathways. Network analysis identified a module primarily containing acute-phase proteins. This module showed lower protein expression in DailyColors™-supplemented participants, strongly corroborating the differential pathway analysis findings. These results suggest a potential anti-inflammatory effect of DailyColors™, consistent with the well-documented effects of the Mediterranean diet. Adherence to the Mediterranean Diet has been associated with reduced systemic inflammation, even in the absence of change in body weight [66–68]. A 3 month study of Mediterranean Diet supplemented with almonds in obese women found a reduction in circulating IL-6 and IL-1ß, as well as an increase in anti-inflammatory M2 macrophage infiltration into subcutaneous adipose tissue [69]. In an 18-month weight loss trial with 294 obese participants, individuals were randomized into three groups: Mediterranean (MED) diet, green-MED diet (enhanced with polyphenols from green tea and polyphenol-rich shakes), and a control group receiving general health advice [70]. Both MED diets led to greater weight loss (MED: −2.7%, green-MED: −3.9%) compared to the control group (−0.4%). At 6 months, the MED and green-MED diets significantly improved cardiovascular and proinflammatory protein profiles, with the green-MED diet showing greater reductions in proteins like IL-18, IL-1 receptor antagonist, and leptin, indicating stronger anti-inflammatory effects. These changes were adjusted for weight loss, though weight reduction may still confound the outcomes. By 18 months, proteomic changes were linked to visceral fat reduction, particularly in the green-MED group, highlighting potential long-term cardiometabolic benefits. A notable strength of this study was its proteomic profiling, although limited to 90 proteins, which may not fully capture all inflammatory pathways. Taken in tandem with our proteomics results, this suggests that DailyColors™ supplementation mimics many of the Mediterranean Diet effects, and if consumed over a longer period of time DailyColors™ supplementation may elicit similar benefits. This requires further study.

The downregulated HDL assembly Reactome pathway described in our results delineates the formation of nascent (discoidal) HDL particles on newly synthesized apoA-I, a process predominantly occurring in the liver. In circulation, these discoidal HDL particles are further lipidated through interactions with cells high in cholesterol, transforming them into spherical HDL. Notably, Sasahara et al. described elevated pre-β1-HDL levels in obese subjects, with BMI positively correlating with pre-β1-HDL and inversely with α1-HDL in both univariate and stepwise regression analyses [71]. Elevated pre-β1-HDL levels in obesity are likely due to increased activities of hepatic lipase and cholesteryl ester transfer protein observed in obese versus lean individuals-HDL particles; this may decrease α1-HDL and elevate pre-β1-HDL levels [72,73]. Therefore, our observation of downregulation of this pathway represents a DailyColors™ induced ‘normalisation’ of a known obesity-induced phenomenon. This is consistent with widespread observational and interventional findings that the Mediterranean diet increases HDL and improves indices of HDL quality [74–76].

There was no effect of DailyColors™ supplementation on concentration or size of total EV concentration or specific EV populations (endothelial- and leukocyte-derived CD31^+^, platelet-derived CD41^+^, leukocyte-derived CD45^+^, or endothelial-derived CD31^+^CD45^-^). Serum proteomic analysis revealed that DailyColors™ elicited decreases in proteins related to vesicle-mediated transport. This Reactome pathway relates broadly to the transport of proteins and other cargo via vesicles, and includes processes such as vesicle formation, coating, budding, uncoating, and fusion with the target membrane. This proteomic observation is in keeping with past findings whereby Mediterranean diet and cocoa flavanol supplementation decreased EV numbers [38,77,78]. However, this was not borne out by EV measurements in the present study, perhaps due to the limited sample size. Positive correlations were identified between EV numbers and plasma 4HBA and hippurate levels. These positive correlations between EV numbers and two of the measured phenolic metabolites suggest that phenolic acids might regulate the formation and release of EVs into the circulation and/or that phenolic acids may be transported within EVs. Indeed, it is known that phenolics can regulate EV biogenesis [79]. Phenolic compound encapsulation within EVs is well-described in plants, cells treated with phenolics are known to alter their cytokine and microRNA cargo and polyphenols can be encapsulated in EVs [79]. What remains to be established is the extent of native phenolic encapsulation in and transport via EVs *in vivo*. Concentrations of platelet-derived (CD41+) and leukocyte-derived (CD45+) EVs were associated with higher BMI; this is concordant with the literature [80]. Concentrations of these EV subclasses were also associated with slower reaction times in picture recognition tasks; higher BMI may be an important confounder as this has been associated with poorer cognitive function [81,82]. Larger mean EV size was associated with improved physical performance and balance, alongside a lower BMI. Increased circulating EV size can be associated with poor cardiometabolic risk profile [83]. Acute physical activity is typically associated with reduced EV size (in men only) [84]. However, obese youths who respond to resistance training by displaying improved insulin sensitivity have larger plasma EVs than non-responders [85]. This finding requires further study.

## Limitations

These data were statistically analysed as within-group pre-post changes. There are several reasons for this. Participants taking the microcrystalline cellulose placebo reported the highest rate of side effects, with 13 individual adverse events reported, of which three were gastrointestinal disturbances, compared with three and one event in the low and high dose respectively of which one was gastrointestinal. This suggests that the placebo may not have been inert and likely interacted with intestinal microbiota, leading to gastrointestinal side effects. The lower-dose DailyColors™ intervention was combined with microcrystalline cellulose as a filler. Our findings indicate a potential functional interaction between these ingredients. Specifically, the low-dose DailyColors™ supplement, but not the high-dose version, led to an increase in circulating isoferulic acid levels two hours after supplementation on day 60. The low-dose supplement also produced the highest circulating concentrations of protocatechuic acid at day 60, although this was not statistically significant. Cellulose and polyphenols are both catabolised by and can in turn alter the intestinal microbiota. Food polyphenols are also known to adsorb to cellulose [86]. Cellulose fermentation with human faeces and without the addition of any polyphenols can elicit the production of phenolic metabolites [87]. It is therefore plausible that this ‘low dose’ phenomenon could be attributed to an interaction between the gut microbiota and the cellulose placebo. However, given that circulating metabolites were measured 2h following capsule ingestion, it is highly improbable that microcrystalline cellulose could have reached the large intestinal microbiome. We hypothesise that cellulose may be exerting effects in the small intestine, and possibly influencing the microbiota there. Despite the prevailing research focus on cellulose’s effects in the colon, there is evidence that a portion of cellulose may be absorbed from the small intestine. Studies with ^14^C-labeled cellulose show a peak in ^14^C-CO₂ expiration at approximately 2 hours post-ingestion, suggesting cellulose presence and absorption in the small intestine [88]. Finally, the similarity in proteomic results observed for both the low and high doses of DailyColors™ suggests that using a pre-post analysis approach with parallel groups is a suitable and effective method for evaluating these interventions.

The relatively short 60-day study duration limits the ability to assess the long-term cognitive and physiological effects of DailyColors™ supplementation. Additionally, the smaller sample size in the subgroup analyses of physiological data reduce statistical power. The primary outcome measure for this pilot study was pathway (not individual protein) level changes in the serum proteome. Hypothesis-free proteomic pathway analyses are not readily conducive to traditional sample size calculations, but our previous work suggested that a subgroup sample size of 15 would be sufficient to identify pathway-level changes [58]. Future research with extended follow-up periods and larger, more diverse samples is required to confirm and expand on these findings. Longer-term studies could help determine whether the observed cognitive benefits are sustained or progress with continued use. Further investigations should be powered *a priori* to explore the mechanistic pathways through which DailyColors™ may act on inflammatory and metabolic processes.

## Conclusion

In conclusion, this study suggests that DailyColors™ supplementation, particularly at higher doses, may support attentional, physical, and systemic health in older overweight adults. The significant improvements in cognitive and physical measures, alongside promising changes in immune-related and HDL protein pathways are consistent with what is known about the effects of the Mediterranean Diet. These changes indicate that DailyColors™ holds potential as a safe, non-pharmacological approach to bolster health and wellbeing in later life. This requires confirmation in larger randomised controlled trials.

## Author contributions

MOL contributed to funding acquisition, conceptualisation, data curation, formal analysis, methodology, project administration, supervision and writing.

JB contributed to funding acquisition, conceptualisation, data curation, formal analysis, methodology, project administration, supervision and writing.

MR contributed to data curation, methodology, project administration, supervision and writing

EB contributed to methodology, formal analysis, project administration and writing

AP contributed to data curation, formal analysis, methodology and writing

KS contributed to project administration and writing

MM contributed to project administration and writing

AB contributed to software, data curation and writing

CB contributed to conceptualisation, formal analysis and writing

AC contributed to funding acquisition, conceptualisation, data curation, formal analysis, methodology, project administration, supervision and writing.

ARM and ZZ conducted the (poly)phenol analysis of the supplements

JCYT conducted analysis of plasma for phenolic metabolites.

## Conflicts of interest

The authors have no conflicts to declare

## Funding Statement

This research was funded by an industrial grant from DailyColors™. The funder had no role in the study design, data collection, analysis, interpretation of data, or decision to publish the results.

## Data Availability Statement

The data supporting the findings of this study are available from the corresponding author upon reasonable request. De-identified datasets will be made publicly accessible in an open-access repository upon acceptance of the manuscript in a peer-reviewed journal.

## Acknowledgements

We thank Dr Kate Heesom (University of Bristol) for her technical support of the proteomics work presented here.

## Notes

### Competing Interest Statement

The authors have declared no competing interest.

### Clinical Trial

ISRCTN10734674

### Author Declarations

Ethics committee/IRB of The University of Exeter Department of Public Health and Sport Sciences gave ethical approval for this work. Ref: 2929219

