## Supplementary Information for "Effects of the DailyColors™ Polyphenol Supplement on Serum Proteome, Cognitive Function, and Health in Older Adults at Risk of Cognitive and Functional Decline"

**Supplementary Table 1**: Capsules and contents provided for each condition, per capsule amounts are provided and participants consumed 4 capsules per day.

| Content | Placebo (mg) | Low Dose (mg) | High Dose (mg) |
|---|---|---|---|
| Chlorophyllin capsule | 123 | 123 | 123 |
| Microcrystalline cellulose | 465 | 357.5 | 65 |
| DailyColors$^{TM}$ Blend | 0 | 187.5 | 500 |
| Natural rice Concentrate | 5 | 5 | 5 |
| Natural rice extract | 5 | 5 | 5 |

**Methods for Analysis of Supplements for Procyanidin, Anthocyanin, Phenolic acids and other (poly)phenols and Flavonols**

**Procyanidin Analysis**

The identification and quantification of flavan-3-ols (monomers and procyanidins) were performed using the method described previously with some modifications (Rodriguez-Mateos et al., 2012; Bussy et al., 2020). Briefly, 2 capsules (1.0 g) of powder were extracted 3 times with a total of 25 mL of acetone / water / acetic acid (70:29.5:0.5, v/v), respectively. Then, samples were vortexed for 5 min, sonicated for 10 min at 50°C, and vortexed again for 2 min, following by centrifugation at 1800 g for 15 min. The supernatants were combined and purified using a Strata SCX SPE cartridge (Strata SCX 500 mg / 3 mL, 55 µm, 70 Å; Phenomenex, Macclesfield, UK), which was pre-conditioned with 3 mL deionised water. The first 1 mL analytical solution was eluted and discarded, while the remaining 2 mL analytical solution was reloaded on the cartridge and the eluent was collected. The collected eluent was filtered through 0.22 µm filters before transferring to HPLC amber vials.

The separation of flavan-3-ol oligomers was performed by an Agilent 1100 series HPLC system (Agilent Technologies, Cheshire, UK) equipped with a fluorescence detector and a Torus Diol column (100 × 3.0 mm, 130 Å, 5.0 µm particle size; Waters UK, Elstree, UK). The separation was accomplished at 40 °C with the injection volume of 5 µL. The mobile phase A was acetonitrile / acetic acid (98:2, v/v) and mobile phase B was methanol / water / acetic acid (95:3:2, v/v). The binary gradients were as follow: 0–1 min, 0% B; 1–5 min, 0-5% B; 5–10 min, 5–50% B; 10–12 min, 50-95% B; 12-15 min, 95% B; 15-17min, 95-0% B, with a flow rate of 0.8 mL/min. The eluate was monitored at 280 nm for UV data, and fluorescence detection was conducted with an excitation of 230 nm and emission at 321 nm.

**Anthocyanin Analysis**

Briefly, 2 capsule powders (1.0 g) were extracted 3 times with a total of 25 mL of acidified methanol (1 % FA, v/v). Samples were vortexed for 5 min, sonicated for 15 min, and centrifuged at 1800 g for 15 min. The supernatant was combined and 5 ml of which was diluted 2-fold with 4% phosphoric acid (v/v) and loaded onto an Oasis PRiME-HLB SPE cartridge. Samples were washed with acidified deionised water (1 % FA, v/v) and isocratic eluted using acidified methanol (1 % FA, v/v) and transferred to HPLC amber vials.

The quantification of anthocyanins in the samples and standard mixes was achieved with a UPLC system (Vanquish, Thermo Fisher Scientific, Runcorn, UK) equipped with electrospray ionisation and triple quadrupole mass spectrometry (Vantage, Thermo Fisher Scientific, Runcorn, UK). Five microliters of samples or standard mixes were injected into the system and passed through a Poroshell 120 SB-C18 column (100 × 3.0 mm, 2.7 μm particle size; Agilent Technologies, Cheshire, UK) with a compatible SB-C18 guard cartridge (5 x 3.0 mm, 2.7 um) and the column temperature of 50 °C. The mobile phases were water / formic acid (98:2, v/v) and methanol / formic acid (99:1, v/v) as solvent A and B, respectively. The gradient were as follows: 0–3 min, 8-18% B; 3–17 min, 18–27% B; 17–18 min, 27–50% B; 18–19 min, 50–95% B; 19–21 min, 95% B; 21-22 min, 95-8%B; and 22-25min, 8% B, with a flow rate of 0.5 mL/min. The MS analysis was performed under the following conditions: collision gas pressure 1.0 mTorr, capillary temperature 350 °C, vaporizer temperature 350 °C, sheath gas pressure 40 arb, aux gas pressure 10 arb, ion sweep gas pressure 2.0 arb, spray voltage 2000. All compounds were analysed in positive ion mode.

Eighteen HPLC grade anthocyanin standards were mixed as the master calibration mix standard, and 13 dilutions were produced for plotting the linear calibration curve. Pelargonidin-3-glucoside (26.42 μmol/L) was fortified into all samples and calibration mixes as the internal standards. The LC-MS parameters of those compounds that lack of commercial standards were firstly obtained from the published references (Mullen et al., 2002; Lopes-da-Sliva et al., 2002; Ludwig et al., 2015; Pertuzatti et al., 2016; Stein-Chisholm et al., 2017; Wang et al., 2019; Wang et al., 2022) and were later verified and modified according to the results of automated optimization on the full sample pool. The final MRM analysis method was generated with the confirmed retention time of the compounds, 2 min RT window, 0.70 FWHM Q1 peak width, and 3.0 seconds cycle time. Peak integration was conducted with the TraceFinder 5.0 Software (Thermo Fisher Scientific, Runcorn, UK) and data calculation was performed on Microsoft Excel. Area ratios of the target compounds to internal standard were used in the quantification to balance the variations in device performance during the batch run.

**Phenolic acids and other (poly)phenol analysis**

Phenolic acids and other (poly)phenol in samples were extracted and quantified according to previous methodologies with some modifications (Dominguez et al., 2021). Briefly, the capsule powder was extracted 3 times with a total of 25 mL of acidified methanol (0.1 % FA, v/v). The same processing method described in the analysis of anthocyanins was applied here for phenolic acids and other (poly)phenols, including vortexing, sonication, centrifugation, and purification by Oasis PRiME-HLB SPE cartridge. However, the isocratic elution solution used here was acidified methanol (0.1 % FA, v/v).

The quantification of phenolic acids and other (poly)phenols in the samples and standard mixes was achieved by the same UPLC-QqQ-MS/MS equipment in anthocyanins analysis. Five microliters of samples or standard mixes were injected into the system and passed through a Raptor Biphenyl reversed phase column 2.1 x 50 mm, 1.8 um (Restek, Bellefonte, USA) coupled with a compatible guard cartridge 5 x 2.1 mm, 2.7 um (Restek, Bellefonte, USA) with the column temperature of 30 °C. The mobile phases were HPLC grade water and HPLC grade acetonitrile that both were acidified with 0.1% LC-MS grade formic acid (v/v), as solvent A and B, respectively. The gradients were as followed: 0–1 min, 1% B; 1–4 min, 1–12% B; 4–8 min, 12% B; 8–11 min, 12–15% B; 11–11.5 min, 15-30% B; 11.5-12 min, 30-99%B; 12-14 min, 99%B; 14-14.1 min 99-1% B; and 14.1-16min, 1% B, with a flow rate of 0.35 mL/min. The MS analysis was performed under the following conditions: collision gas pressure 1.0 mTorr, capillary temperature 270 °C, vaporizer temperature 350 °C, sheath gas pressure 49 arb, aux gas pressure 10 arb, ion sweep gas pressure 0.0 arb, spray voltage 3000. All of the compounds were analysed in negative ion mode.

Forty six HPLC grade standards were used to identify and quantify phenolic acids and other (poly)phenols. All the standards were mixed as the master calibration mix and 13 dilutions were made for plotting the calibration curve. Taxifolin (0.25 mg/mL) was fortified into all samples and calibration mixes as the internal standard. Then, the final MRM analysis method was generated accordingly with the confirmed retention time of the compounds, 2 min RT window, 0.70 FWHM Q1 peak width, and 3.0 seconds cycle time. The peak integration was conducted with the TraceFinder 5.0 Software (Thermo Fisher Scientific, Runcorn, UK) and data calculation was performed on Microsoft Excel as described in anthocyanins analysis.

**Flavonol Analysis**

After the same extraction process as for phenolic acids, the UPLC-MS analysis of flavonols in the samples and standard mixes was achieved with a UPLC system (Thermo Fisher Scientific,

Runcorn, UK) equipped with electrospray ionisation and triple quadrupole mass spectrometry (Quantum access, Thermo Fisher Scientific, Runcorn, UK). Five microliters of samples or standard mixes were injected into the system and passed through a Poroshell 120 EC-C18 column (100 × 2.1 mm, 2.7 μm particle size; Agilent Technologies, Cheshire, UK) with a compatible EC-C18 guard cartridge 5 x 2.1 mm, 2.7 um and the column temperature of 25 °C. The mobile phases were HPLC grade water and acetonitrile that both were acidified with 0.1% formic acid (v/v), as solvent A and B, respectively. The gradients were as followed: 0–0.2 min, 5% B; 0.2–2 min, 5–17% B; 2–10 min, 17–19.5% B; 10–10.5 min, 19.5-50% B; 10.5–11 min, 50-90% B; 11-13 min, 90%B; 13-13.5 min, 90-5%B; and 13.5-16min, 5% B, with a flow rate of 0.4 mL/min by the ACCELA quaternary pump. The MS analysis was performed under the following conditions: collision gas pressure 1.5 mTorr, capillary temperature 300 °C, vaporizer temperature 350 °C, sheath gas pressure 40 arb, aux gas pressure 10 arb, ion sweep gas pressure 0.0 arb, spray voltage 2500. All of the compounds were analysed in negative ion mode.

Nine HPLC grade standards were used for the identification and quantification of the flavonols. The master calibration standard mix was made, and 8 dilutions were produced for plotting the calibration curve. Taxifolin (0.25 mg/mL) was fortified into all samples and calibration mixes as the internal standard. The LC-MS parameters of those compounds that lack of commercial standards were firstly obtained from the published references (Vrhovsek et al., 2012; Alvarez-Fernandez et al., 2015; Pertuzatti et al., 2021; Renai et al., 2021) and were later verified and modified according to the results of automated optimization on the full sample pool. The final MRM analysis method was generated accordingly with the confirmed RT, 2 min RT window, 0.70 FWHM Q1 and Q3 peak width, 0.2 m/z scan width and 0.005 s scan time. The peak integration was conducted with the Xcalibur Quan Browser 4.3.73.11 Software (Thermo Fisher Scientific, Runcorn, UK) and data calculation was performed on Microsoft Excel as described in anthocyanins analysis.

**Supplementary Table 2.** Daily dose of measured (poly)phenols provided by consumption of 4 capsules per day, and composition of proprietary DailyColors$^{TM}$ blend.

| Compound | Placebo | Low dose | High dose | DC Blend (mg/100g) |
|---|---|---|---|---|
| **Anthocyanins (mg/d)** | 0.000 | 34.226 | 93.695 | 5884.933 |
| **Flavonols (mg/d)** | 0.033 | 5.365 | 9.262 | 522.353 |
| **Phenolic acids (mg/d)** | 0.065 | 66.417 | 109.265 | 5343.034 |
| **Flavan-3-ols (mg/d)** | 1.044 | 87.255 | 180.136 | 9741.573 |
| **Other (Poly)Phenols (mg/d)** | 0.473 | 53.392 | 69.799 | 3460.106 |
| **Total measured (Poly)Phenol content (mg/d)** | 1.615 | 246.655 | 462.157 | 24951.999 |

**Supplementary Table 3.** Daily dose of phenolic compounds provided by consumption of 4 capsules per day, and composition of proprietary DailyColors™ blend.

| Compounds | Placebo (mg/d) | Low dose (mg/d) | High Dose (mg/d) | DC Blend (mg/100g) |
|---|---|---|---|---|
| **Anthocyanins** | | | | |
| Cyanidin-3-aldopentoside | 0.000 | 0.040 | 0.111 | 1.055 |
| Cyanidin-3-arabinoside | 0.000 | 0.479 | 1.413 | 7.306 |
| Cyanidin-3-galactoside | 0.000 | 0.917 | 2.320 | 88.509 |
| Cyanidin-3-glucoside | 0.000 | 1.673 | 4.517 | 130.072 |
| Cyanidin-3-glucoside-chalcone | 0.000 | 0.009 | 0.022 | 288.265 |
| Cyanidin-3-glucuronide | 0.000 | 0.090 | 0.262 | 16.266 |
| Cyanidin-3-hexoside | 0.000 | 0.072 | 0.265 | 16.599 |
| Cyanidin-3-rutinoside | 0.000 | 0.957 | 2.618 | 182.149 |
| Delphinidin-3-arabinoside | 0.000 | 1.691 | 4.632 | 283.345 |
| Delphinidin-3-galactoside | 0.000 | 3.448 | 8.767 | 609.955 |
| Delphinidin-3-glucoside | 0.000 | 7.672 | 20.997 | 1208.819 |
| Delphinidin-3-glucoside-chalcone | 0.000 | 0.063 | 0.116 | 6.648 |
| Delphinidin-3-hexoside 1 | 0.000 | 0.076 | 0.155 | 9.896 |
| Delphinidin-3-hexoside 2 | 0.000 | 0.042 | 0.084 | 5.270 |
| Delphinidin-3-xyloside | 0.000 | 0.453 | 1.092 | 66.538 |
| Malvidin-3-arabinoside | 0.000 | 4.398 | 12.208 | 784.972 |
| Malvidin-3-dihexoside | 0.000 | 0.018 | 0.041 | 2.370 |
| Malvidin-3-galactoside | 0.000 | 0.881 | 2.609 | 161.085 |
| Malvidin-3-glucoside | 0.000 | 5.014 | 13.259 | 825.693 |
| Pelargonidin-3-glucoside | 0.000 | 0.030 | 0.078 | 4.691 |
| Peonidin-3-arabinoside | 0.000 | 0.090 | 0.212 | 14.316 |
| Peonidin-3-galactoside | 0.000 | 0.307 | 0.761 | 56.611 |
| Peonidin-3-glucoside | 0.000 | 0.652 | 1.689 | 103.122 |
| Peonidin-3-glucoside-chalcone | 0.000 | 0.530 | 1.530 | 94.081 |
| Petunidin-3-arabinoside | 0.000 | 1.139 | 2.766 | 176.048 |
| Petunidin-3-galactoside | 0.000 | 1.580 | 5.852 | 426.312 |
| Petunidin-3-glucoside | 0.000 | 1.905 | 5.319 | 314.940 |
| **Total** | **0.000** | **34.226** | **93.695** | **5884.933** |
| **Flavonols** | | | | |
| Quercetin-3-rhamnoside-1 | 0.000 | 0.064 | 0.142 | 9.139 |

| Compounds | Placebo (mg/d) | Low dose (mg/d) | High Dose (mg/d) | DC Blend (mg/100g) |
|---|---|---|---|---|
| Quercetin-3-rhamnoside-2 | 0.000 | 0.005 | 0.006 | 0.215 |
| Quercetin-3-rhamnoside-3 | 0.002 | 0.011 | 0.010 | 0.512 |
| Quercetin-3-hexoside-1 | 0.002 | 0.092 | 0.086 | 3.696 |
| Quercetin-3-hexoside-2 | 0.000 | 0.184 | 0.358 | 18.148 |
| Quercetin-3-hexoside-3 | 0.000 | 0.601 | 0.961 | 57.490 |
| Quercetin-3-hexoside-4 | 0.002 | 0.040 | 0.050 | 2.710 |
| Quercetin-3-aldopentoside-1 | 0.000 | 0.059 | 0.064 | 2.969 |
| Quercetin-3-aldopentoside-2 | 0.000 | 0.056 | 0.062 | 2.842 |
| Quercetin-3-aldopentoside-3 | 0.000 | 0.184 | 0.270 | 15.242 |
| Quercetin-3-rutinoside | 0.000 | 0.002 | 0.010 | 0.519 |
| Quercetin-3-acetyl-hexoside | 0.017 | 1.210 | 2.273 | 120.372 |
| Quercetin-3-deoxyhexose-hexoside | 0.000 | 0.014 | 0.036 | 1.844 |
| Kaempferol-3-aldopentoside | 0.000 | 0.047 | 0.051 | 2.851 |
| Kaempferol-3-hexoside-1 | 0.000 | 0.091 | 0.106 | 4.594 |
| Kaempferol-3-hexoside-2 | 0.000 | 0.645 | 1.073 | 60.217 |
| Kaempferol-3-rutinoside-1 | 0.000 | 0.021 | 0.047 | 2.656 |
| Kaempferol-3-rutinoside-2 | 0.000 | 0.029 | 0.026 | 1.441 |
| Kaempferol-3-rutinoside-3 | 0.003 | 0.067 | 0.149 | 9.184 |
| Isorhamnetin-3-glucoside | 0.000 | 0.012 | 0.023 | 1.423 |
| Isorhamnetin-3-hexoside-1 | 0.000 | 0.051 | 0.052 | 2.519 |
| Isorhamnetin-3-hexoside-2 | 0.000 | 0.051 | 0.053 | 2.471 |
| Isorhamnetin-3-aldopentoside | 0.000 | 0.006 | 0.007 | 0.493 |
| Isorhamnetin-3-rutinoside | 0.000 | 0.012 | 0.018 | 1.142 |
| Isorhamnetin-3-acetyl-hexoside | 0.000 | 0.001 | 0.003 | 0.184 |
| Isorhamnetin-3-(6-rhamnosyl)-galactoside | 0.000 | 0.032 | 0.070 | 4.418 |
| Laricitrin-3-hexoside | 0.000 | 0.157 | 0.272 | 16.613 |
| Laricitrin-3-rhamnoside | 0.002 | 1.226 | 2.152 | 122.437 |
| Myricetin-3-rhamnoside | 0.005 | 0.007 | 0.023 | 1.280 |
| Myricetin-3-aldopentoside | 0.000 | 0.276 | 0.610 | 40.695 |
| Myricetin-3-hexoside | 0.000 | 0.078 | 0.139 | 7.716 |
| Syringetin-3-hexoside | 0.000 | 0.034 | 0.060 | 4.321 |

| Compounds | Placebo (mg/d) | Low dose (mg/d) | High Dose (mg/d) | DC Blend (mg/100g) |
|---|---|---|---|---|
| **Total** | **0.033** | **5.365** | **9.262** | **522.353** |
| **Phenolic Acids** | | | | |
| 2,3-Dihydroxybenzoic acid | 0.000 | 2.979 | 5.700 | 267.533 |
| 2,4-Dihydroxybenzoic acid | 0.000 | 0.000 | 0.000 | 0.023 |
| 2,5-Dihydroxybenzoic acid | 0.000 | 0.058 | 0.104 | 5.474 |
| 2,6-Dihydroxybenzoic acid | 0.000 | 0.000 | 0.000 | 0.002 |
| 3-Feruloylquinic acid | 0.000 | 1.566 | 2.837 | 133.183 |
| 3-methylgallic acid | 0.000 | 0.006 | 0.011 | 0.876 |
| 3,4-dihidroxyphenylacetic acid | 0.000 | 1.859 | 3.480 | 156.040 |
| 3,4-dihydroxycinnamic acid | 0.000 | 20.351 | 34.174 | 1708.093 |
| 3,5-Dihydroxybenzoic acid | 0.000 | 4.062 | 7.770 | 364.668 |
| 4-Caffeoylquinic acid | 0.000 | 1.641 | 2.595 | 122.436 |
| 4-Feruloylquinic acid | 0.000 | 5.116 | 10.177 | 502.640 |
| 4-Hydroxybenzoic acid | 0.000 | 0.595 | 1.123 | 53.321 |
| 4-methoxysalicylic acid | 0.000 | 0.003 | 0.006 | 0.486 |
| 6-methoxysalicylic acid | 0.000 | 0.013 | 0.023 | 1.280 |
| Caffeic acid | 0.000 | 5.718 | 7.602 | 364.113 |
| Caffeic acid hexoside | 0.000 | 0.009 | 0.011 | 0.586 |
| Caffeic acid derivative | 0.000 | 0.049 | 0.073 | 3.766 |
| Chlorogenic acid | 0.005 | 14.733 | 20.931 | 1074.336 |
| Feruloyl-tartaric acid | 0.001 | 0.001 | 0.002 | 0.038 |
| Gallic acid | 0.000 | 6.235 | 10.397 | 472.683 |
| Homo Protocatechuic acid | 0.000 | 0.000 | 0.000 | 0.015 |
| Homo Vanillic acid | 0.000 | 0.062 | 0.131 | 9.514 |
| Isoferulic acid | 0.029 | 0.000 | 0.327 | 14.042 |
| Isovanillic acid | 0.000 | 0.002 | 0.005 | 0.340 |
| L-Tartaric acid | 0.015 | 0.941 | 1.074 | 54.486 |
| p-coumaric acid | 0.012 | 0.176 | 0.296 | 14.407 |
| Protocatechuic acid | 0.000 | 0.000 | 0.001 | 0.033 |
| Sinapic acid | 0.000 | 0.004 | 0.005 | 0.232 |
| Syringic acid | 0.003 | 0.182 | 0.263 | 12.331 |
| trans-ferulic acid | 0.000 | 0.056 | 0.147 | 6.057 |
| **Total** | **0.065** | **66.417** | **109.265** | **5343.034** |

| Compounds | Placebo (mg/d) | Low dose (mg/d) | High Dose (mg/d) | DC Blend (mg/100g) |
|---|---|---|---|---|
| **Flavan-3-ols** | | | | |
| DP1 | 0.216 | 24.639 | 24.810 | 1249.269 |
| DP2 | 0.114 | 19.019 | 42.197 | 2303.264 |
| DP3 | 0.118 | 15.579 | 38.740 | 2020.986 |
| DP4 | 0.113 | 11.269 | 32.745 | 1783.513 |
| DP5 | 0.118 | 7.497 | 18.962 | 1061.483 |
| DP6 | 0.108 | 4.603 | 10.675 | 606.015 |
| DP7 | 0.086 | 2.440 | 6.216 | 413.013 |
| DP8 | 0.085 | 1.044 | 2.834 | 166.021 |
| DP9 | 0.086 | 0.730 | 1.858 | 73.299 |
| DP10 | 0.000 | 0.435 | 1.099 | 64.709 |
| **Total** | **1.044** | **87.255** | **180.136** | **9741.572** |
| **Other (Poly)phenols** | | | | |
| 4-Hydroxybenzaldehyde | 0.090 | 0.077 | 0.179 | 2.312 |
| Gallic acid ethyl ester | 0.000 | 0.006 | 0.005 | 0.267 |
| Genistein | 0.001 | 0.737 | 1.382 | 61.646 |
| Hesperetin | 0.001 | 0.219 | 0.297 | 13.840 |
| Isorhamnetin | 0.004 | 0.611 | 0.753 | 41.511 |
| Kaempferol | 0.034 | 0.350 | 0.439 | 23.908 |
| Laricitrin | 0.000 | 0.007 | 0.012 | 0.665 |
| Luteolin | 0.001 | 0.043 | 0.052 | 2.750 |
| Morin | 0.137 | 25.251 | 32.009 | 1592.536 |
| Myricetin | 0.002 | 0.274 | 0.335 | 15.171 |
| Phloretin | 0.015 | 1.812 | 2.733 | 138.914 |
| Phloretin-2-glucoside | 0.000 | 0.002 | 0.003 | 0.127 |
| Protocatechuic acid methyl ester | 0.000 | 0.000 | 0.000 | 0.051 |
| Pyrogallol | 0.000 | 0.434 | 0.848 | 39.970 |
| Quercetin | 0.156 | 22.175 | 28.111 | 1398.590 |
| Rhamnetin | 0.001 | 0.000 | 0.001 | 0.046 |
| Secoisolariciresinol | 0.031 | 0.057 | 0.108 | 4.261 |
| Tyrosol | 0.000 | 1.240 | 2.345 | 114.092 |
| Vanillin | 0.000 | 0.097 | 0.187 | 9.449 |
| **Total** | **0.473** | **53.392** | **69.799** | **3460.106** |

| Compounds | Placebo (mg/d) | Low dose (mg/d) | High Dose (mg/d) | DC Blend (mg/100g) |
|---|---|---|---|---|
| **Total (Poly)phenol Content** | **1.615** | **246.655** | **462.157** | **24951.998** |

**Plasma Phenolic Metabolite Assay**

In brief, 200 µL of plasma, calibration standards (blank plasma spiked with European Pharmacopoeia (EP) reference standards obtained from Merck Germany), and plasma quality controls were spiked with 20 µL of internal standard containing ferulic acid-[2H3] (100 nmol.L-1) and hippuric acid [13C6] (200 µmol/L) (Toronto Research Chemicals, Ontario, Canada) in 0.1% formic acid (Merck, Germany) into microcentifuge tubes and mixed. To this, 1 mL of methanol was added slowly with gentle mixing, the mixture was then incubated at room temperature for 15 minutes, followed by centrifugation at 14,000 rpm for 7 minutes. The supernatant was transferred to borosilicate glass tube and placed in an evaporator to dry under a constant stream of nitrogen at a temperature of 60°C. To the dried supernatant 200 µL of methanol (Merck, Germany) with 0.1% formic acid was added into each tube and vortex mixed for 30 seconds, followed by 2.5 mL of ethylacetate (Merck, Germany) and vigorously mixed for 10 minutes. After centrifugation at 4,000 rpm for 10 minutes, 2 mL of the ethylacetate in the upper layer was transferred to a fresh set of borosilicate glass tubes and again evaporated to dryness as described above. The dried residue was resuspended in 250 µL of LCMS grade deionised water with 1% acetic acid (Merck, Germany), then vortex mixed followed by centrifugation at 4,000 rpm for 10 mins. The final mixture was transferred into polypropylene autosampler vials, 50 µL was injected into the LC-MS/MS for analysis. MassLynx version 4.2 and QuanLynx software (Waters Corp., Milford, MA, USA) were used for system control, data acquisition, baseline integration and peak quantification.

**Supplementary Table 4: Plasma phenolic metabolite assay characteristics**

| | **Protocate-chuic acid nmol/L** | **4-hydroxy-benzoic acid nmol/L** | **hippuric acid µmol/L** | **vanillic acid nmol/L** | **ferulic acid nmol/L** | **isoferulic acid nmol/L** |
|---|---|---|---|---|---|---|
| Linearity | 0.5-486 | 1.0-749 | 0.01-800 | 0.5-336.7 | 0.5-321.1 | 0.5-337.7 |
| Typical $r^2$ | >0.98 | >0.98 | >0.98 | >0.98 | >0.98 | >0.98 |
| Intra assay imprecision conc. mean (%CV), n=6 | 39.4 (2.6)<br>79.4 (5.8)<br>430.4 (2.2) | 57.3 (3.5)<br>261.6 (2.5)<br>622.9 (2.0) | 6.1 (0.3)<br>34.7 (0.9)<br>499.2 (2.4) | 36.9 (9.5)<br>43.7 (5.9)<br>250.9 (7.6) | 3.3 (6.6)<br>9.4 (3.7)<br>143.9 (2.1) | 2.1 (4.0)<br>106.2 (4.2)<br>218.6 (4.4) |
| Inter assay imprecision conc. mean (%CV), n=6 | 20.0 (10.6)<br>135.1 (9.0)<br>424.7 (7.0) | 32.9 (6.0)<br>393.6 (6.2)<br>622.8 (7.6) | 32.9 (6.0)<br>393.6 (6.2)<br>522.8 (7.6) | 35.7 (5.4)<br>221.6 (6.8)<br>359.4 (9.2) | 4.5 (6.1)<br>132.2 (6.1)<br>294 (7.0) | 3.7 (9.9)<br>133.6 (6.5)<br>277 (7.5) |
| Lower Limits of quantificatio n (LLoQ) | 1.0 | 1.0 | 0.05 | 1.0 | 1.0 | 1.0 |
| Spiked recovery Mean% (±SD)* | 98.5% (±2) | 96.8% (±3) | 103% (±2) | 105% (±2) | 99% (±2) | 102% (±3) |

* Base serum used for spiking contained 100 µmol.L$^{-1}$ of hippuric acid and no other endogenous phenolic metabolites. Each spiked sample was tested six times.

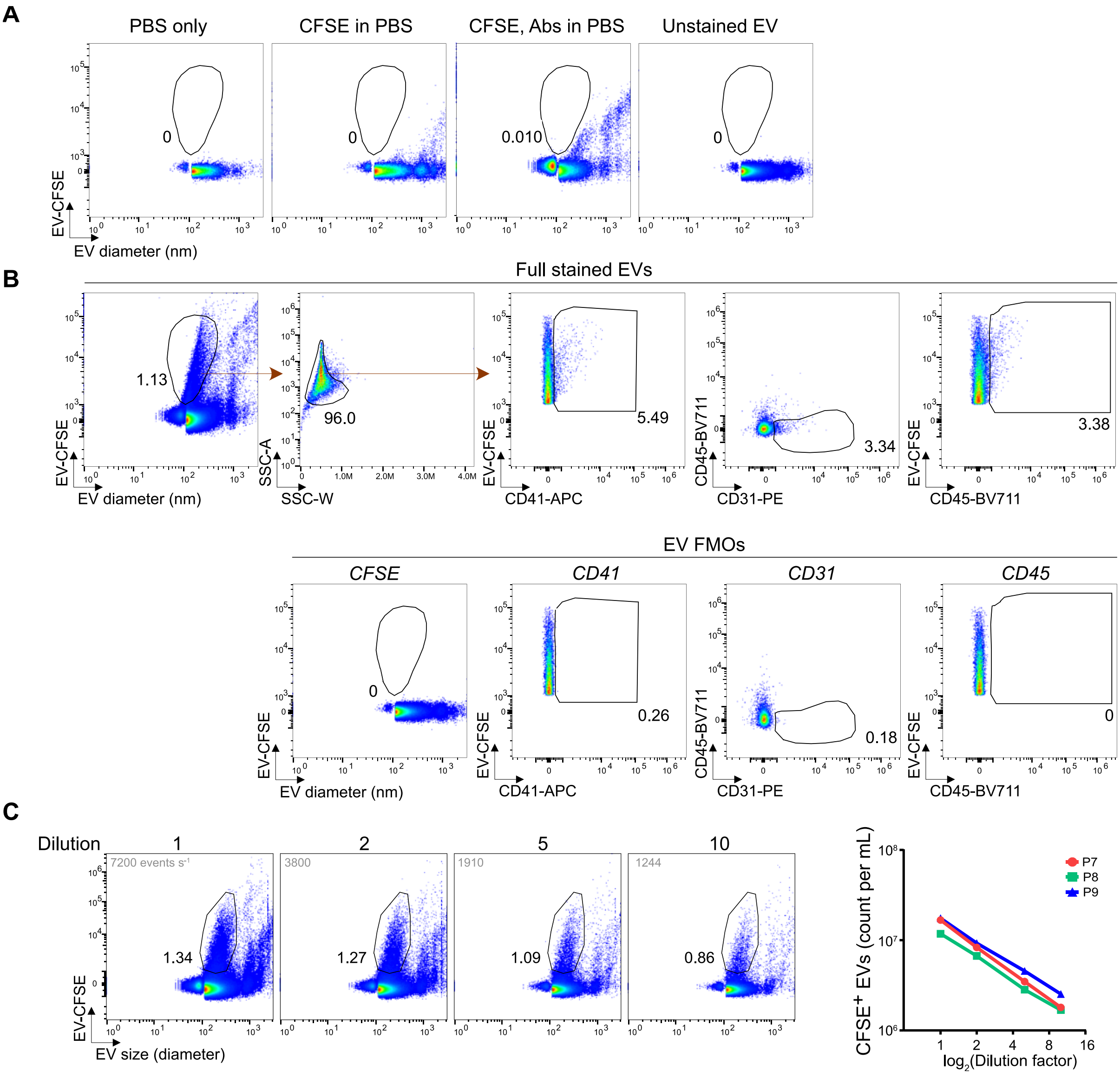


**Supplementary Figure 1.** Flow cytometry gating of EVs. A) Representative flow cytometry plots of PBS alone, CFSE in PBS, a complete staining mixture in PBS or unstained EVs. B) A representative flow cytometry gating strategy demonstrating EV and subpopulation identification using fluorescence minus one (FMO) controls. C) A serial dilution was performed on stained EVs. Flow cytometry plots and graphs demonstrate a proportional decrease EV concentration with dilution.

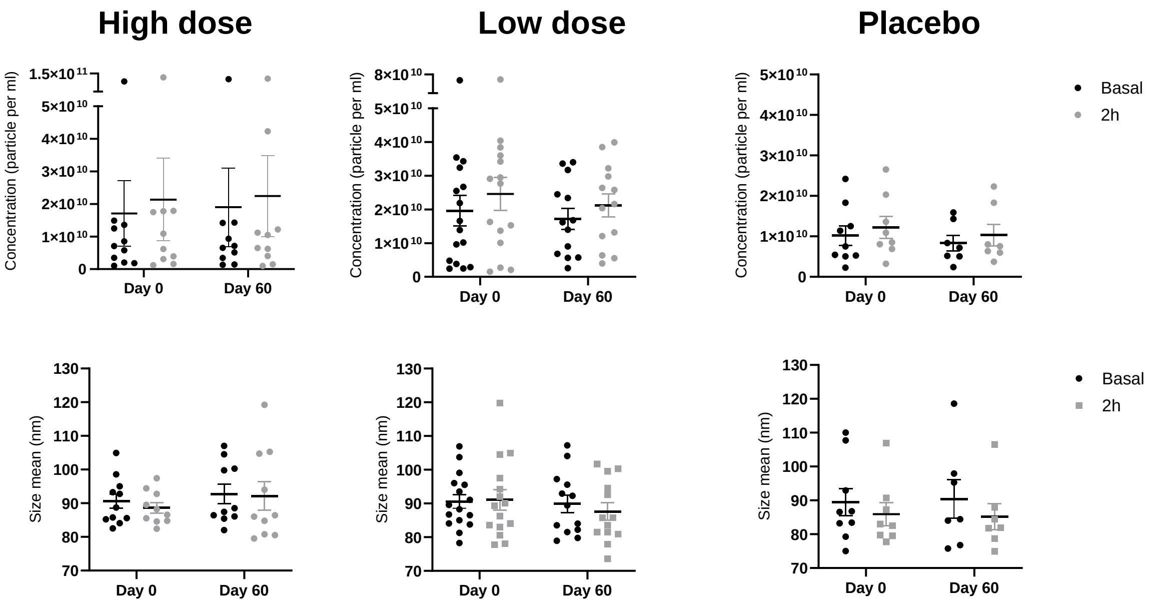


**Supplementary Figure 2.** Sixty days DailyColors™ supplementation does not alter circulating extracellular vesicle number or size, as measured by nanoparticle tracking analysis.

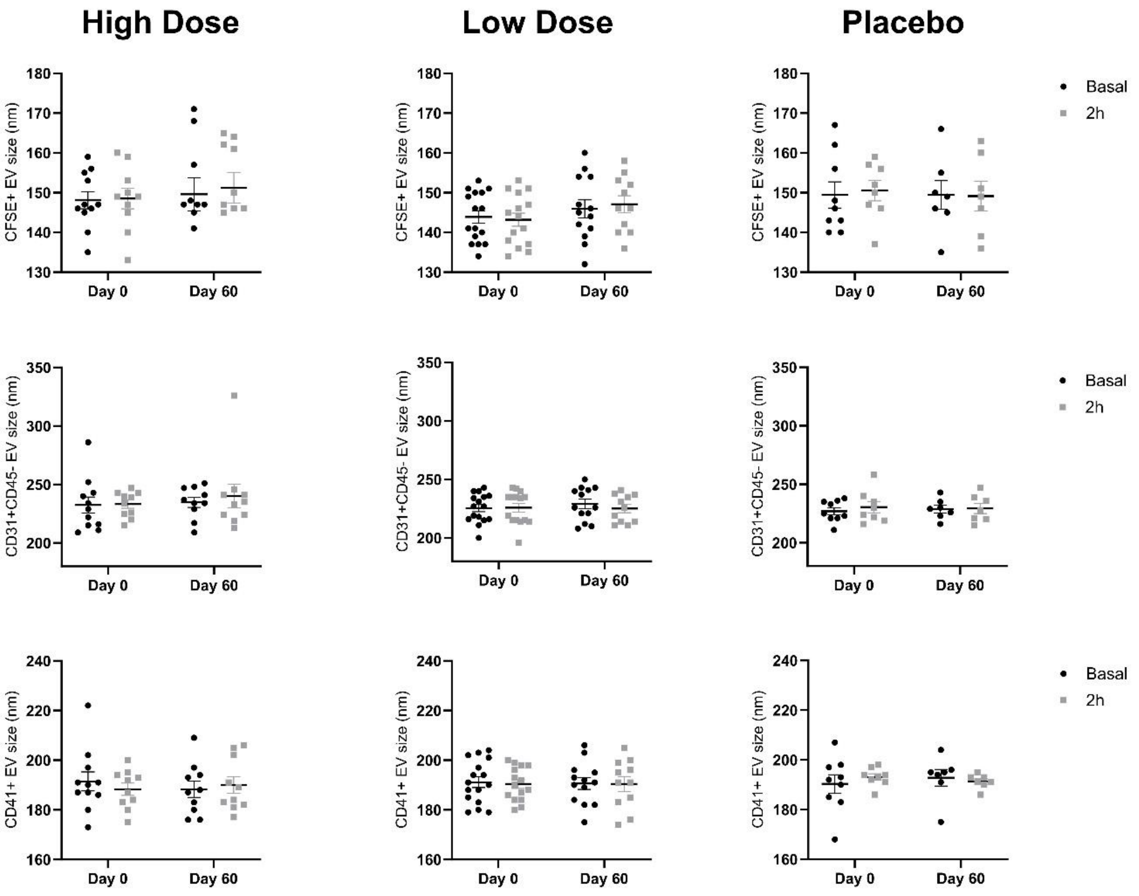


**Supplementary Figure 3.** Sixty days DailyColors™ supplementation does not alter counts of specific EV populations measured via flow cytometry. CFSE = Carboxyfluorescein succinimidyl ester.

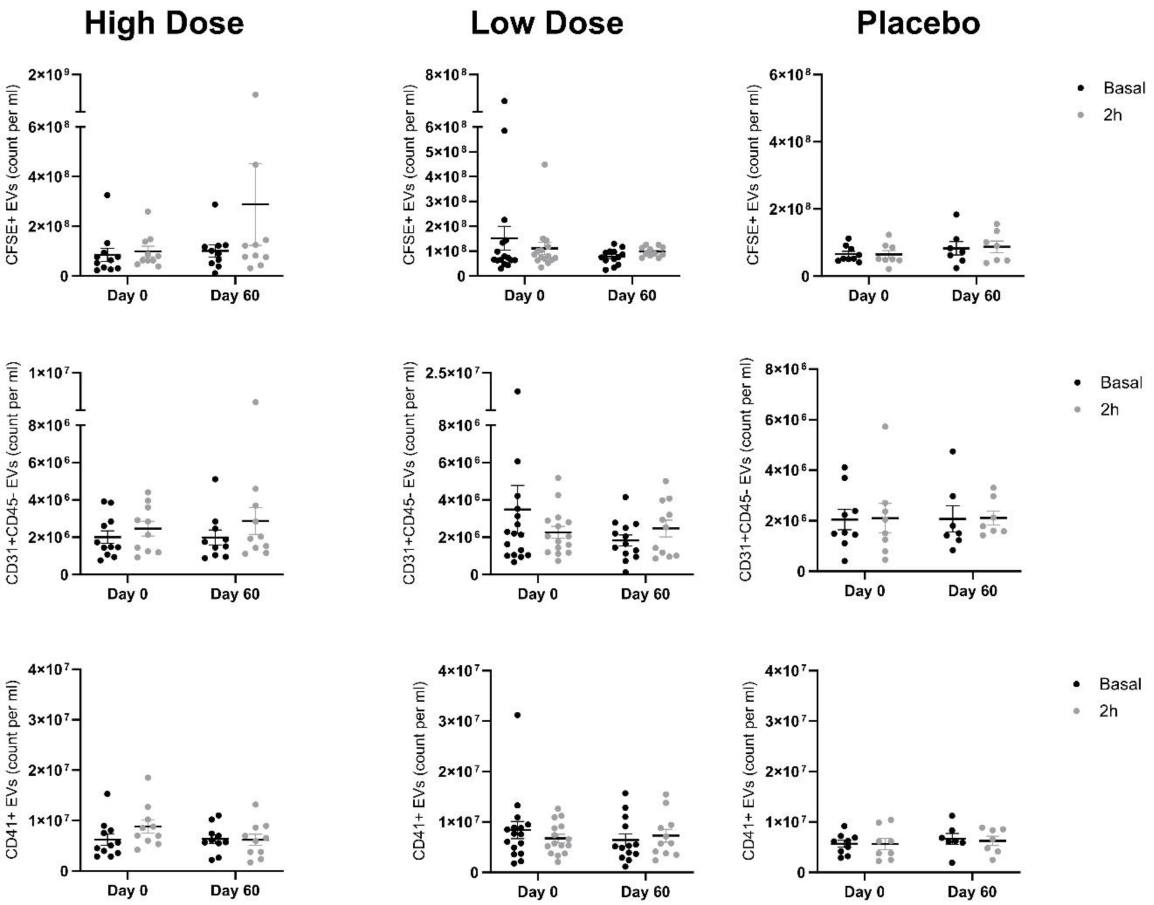


**Supplementary Figure 4.** Sixty days DailyColors™ supplementation does not alter size of specific EV populations measured via flow cytometry. CFSE = Carboxyfluorescein succinimidyl ester.

**Supplementary Table 5: Baseline Characteristics for the trial cohort and treatment groups**

| | Measure | | Whole Cohort | Placebo Group | DailyColors™ Low Dose | DailyColors™ High Dose |
|---|---|---|---|---|---|---|
| | | n | 152 | 51 | 50 | 50 |
| Stratification Variables | Age | Range | 51-86 | 51-78 | 51-86 | 53-83 |
| | | Mean (SD) | 64 (6.7) | 63.5 (6.29) | 64.4 (7.24) | 64.1 (6.64) |
| | Sex | Female (%) | 123 (80) | 41 (80) | 41 (80) | 41 (80) |
| | | Male (%) | 30 (20) | 10 (20) | 10 (20) | 10 (20) |
| | BMI | Range | 25.1 - 60.8 | 25.4 - 60.8 | 25.1 - 38.3 | 25.2 - 52.3 |
| | | Mean (SD) | 30.0 (5.09) | 30.2 (5.54) | 28.7 (3.48) | 31.1 (5.75) |
| Cognition | Choice Reaction Time | Range | 399 - 906 | 422 – 693.5 | 410 - 794 | 454 – 804.5 |
| | | Mean (SD) | 540.00 (66.85) | 545.28 (59.56) | 525.89 (68.66) | 544.75 (62.30) |
| | Digit Vigilance Reaction Time | Range | 389.3 - 723.4 | 394.4 - 654.9 | 389.3 - 723.4 | 408.3 - 664.9 |
| | | Mean (SD) | 490.14 (56.29) | 482.39 (56.01) | 488.82 (60.00) | 494.72 (51.45) |
| | Picture Recognition Accuracy | Range | 50 - 100 | 60 – 100 | 55 - 100 | 70 - 100 |
| | | Mean (SD) | 92.02 (9.17) | 92.4 (8.34) | 92.55 (9.02) | 92.1 (8.52) |
| | Picture Recognition Reaction Time | Range | 701.5 - 1774 | 877.5 - 1549.5 | 777.7 - 1752.1 | 841.4 - 1836.2 |
| | | Mean (SD) | 1101.21 (197.24) | 1150.07 (183.24) | 1288.87 (219.73) | 1305 (232.63) |

| | | | | | | |
|---|---|---|---|---|---|---|
| | **Spatial Working Memory** | Range | 0 - 7 | 2 - 6 | 0 - 7 | 3 - 6 |
| | | Mean (SD) | 4.28 (0.90) | 4.25 (0.86) | 4.25 (1.08) | 4.35 (0.79) |
| | **Executive Function** | Range | 8 - 76 | 12 - 71 | 17 - 71 | 8 - 64 |
| | | Mean (SD) | 38.87 (11.42) | 40.7 (12.46) | 39.12 (9.52) | 37.38 (10.92) |
| **Physical Fitness** | **Chair Stand Test** | Range | 0 – 33 | 0 – 33 | 9 – 28 | 6 – 27 |
| | | Mean (SD) | 15.32 (4.88) | 15.69 (5.44) | 15.78 (5.29) | 14.55 (4.02) |
| | **Timed Stand Test** | Range | 1 – 60 | 1 – 60 | 9 – 29 | 7 – 32 |
| | | Mean (SD) | 19.01 (6.55) | 19.35 (8.18) | 19.31 (5.46) | 19.34 (6.10) |
| | **2-minute Step Test** | Range | 1 – 199 | 1 – 199 | 62 – 134 | 64 – 127 |
| | | Mean (SD) | 92.51 (25.05) | 88.75 (32.77) | 105.65 (20.10) | 95.13 (16.26) |
| | **ABC Scale total score (Balance)** | Range | 1.5 – 5 | 1.5 – 5 | 2.1 – 5 | 3.1 – 5 |
| | | Mean (SD) | 4.63 (0.59) | 4.57 (0.7) | 4.42 (0.73) | 4.61 (0.43) |
| **Wellbeing** | **IQCODE score (Subjective Cognition)** | Range | 1.75 – 3.94 | 2.5 – 3.94 | 3.06 – 3.75 | 3.06 – 3.56 |
| | | Mean (SD) | 3.11 (0.25) | 3.14 (0.23) | 3.24 (0.12) | 3.25 (0.12) |
| | **EQ5D Score (Quality of Life)** | Range | 0.1 – 1 | 0.12 – 1.0 | 0.3 – 1 | 0.42 – 1 |
| | | Mean (SD) | 0.82 (0.16) | 0.80 (0.17) | 0.79 (0.16) | 0.80 (0.13) |
| | **GAD-7 Score (Anxiety)** | Range | 0 – 19 | 0 – 19 | 0 – 16 | 0 – 17 |
| | | Mean (SD) | 2.22 (3.40) | 2.51 (3.45) | 2.94 (3.55) | 2.76 (3.70) |
| | **PHQ-9 Score** | Range | 0 – 23 | 0 – 15 | 0 – 14 | 0 – 23 |

| | | | | | |
|---|---|---|---|---|---|
| **(Depression)** | Mean (SD) | 3.73 (4.01) | 3.58 (3.47) | 4.8 (4.36) | 4.93 (5.22) |

**Supplementary Table 6: Impact of DailyColors supplementation on cognitive, health and wellbeing measures**

| Measure | | Placebo Group | | | DailyColors™ Low Dose | | | DailyColors™ High Dose | | |
|---|---|---|---|---|---|---|---|---|---|---|
| | | Effect Size | CI | P | Effect Size | CI | P | Effect Size | CI | P |
| Cognition | Choice Reaction Time | 0.38 | 0.56 - 0.707 | **<0.001** | 0.11 | -0.431 - 0.207 | 0.490 | 0.19 | -0.508 - 0.133 | 0.252 |
| | Digit Vigilance Reaction Time | 0.19 | -0.13 - 0.503 | 0.248 | 0.005 | -0.309 - 0.319 | 0.975 | 0.34 | 0.012 - 0.667 | **0.042** |
| | Picture Recognition Accuracy | 0.19 | -0.508 - 0.126 | 0.238 | 0.42 | -0.744 - - 0.81 | 0.15 | 0.42 | -0.756 - -0.83 | **0.014** |
| | Picture Recognition Reaction Time | 0.66 | 0.309 - 1.003 | **<0.001** | 0.39 | 0.060 - 0.720 | **0.021** | 0.14 | 0.278 - 0.986 | **<0.001** |
| | Spatial Working Memory | 0.09 | -0.444 - 0.178 | 0.403 | 0.18 | -0.500 - 0.133 | 0.255 | 0.26 | -0.565 - 0.57 | 0.110 |
| | Executive Function | 0.61 | -0.950 0 –0.265 | **<0.001** | 0.05 | -0.267 - 0.369 | 0.755 | 0.20 | -0.522 - 0.129 | 0.238 |
| Physical Fitness & Wellbeing | Chair Stand Test | 0.36 | -0.674 - -0.34 | 0.030 | 0.76 | -1.115 - 0.400 | **<0.001** | 0.50 | -0.146 - 0.406 | **0.001** |
| | Timed Stand Test | 0.03 | -0.222 - 0.399 | 0.575 | 0.20 | -0.116 - 0.518 | 0.213 | 0.67 | -0.344 - 0.222 | **<0.001** |
| | 2-minute Step Test | 0.44 | -0.762 - -0.112 | 0.323 | 0.035 | -0.279 - 0.349 | 0.826 | 0.36 | -0.646 - -0.062 | **0.017** |
| | ABC Scale total score (Balance) | 0.39 | 0.112 - 0.682 | **0.006** | 0.31 | 0.027 - 0.595 | **0.032** | 0.13 | -0.146 - 0.406 | 0.355 |

| | | | | | | | | | | |
|---|---|---|---|---|---|---|---|---|---|---|
| | **PHQ-9 Score (Depression)** | 0.49 | 0.112 - 0.682 | **<0.001** | 0.49 | 0.026 - 0.585 | **0.001** | 0.38 | 0.93 - 0.661 | **0.009** |
